# Psilocybin-Assisted Early Palliative Care for Demoralization and Chronic Pain: An Open-Label Pilot Study

**DOI:** 10.64898/2026.07.11.26357570

**Authors:** Ali J. Zarrabi, Tanja Mletzko, George Grant, Caroline Peacock, Roman Palitsky, Tarrant McPherson, Isabelle Shub, Sarah Eisenacher, Jessica L. Maples-Keller, Deanna Kaplan, Barbara O. Rothbaum, Fayzan Rab, Nupur Dalal, Kimberly A. Curseen, Charles Raison, Boadie W. Dunlop

## Abstract

**Background:** Demoralization, a syndrome of helplessness, hopelessness, and loss of meaning and chronic pain are common sources of distress in early palliative care. Psilocybin-assisted therapy (PAT) is an emerging intervention with preliminary data suggesting improvements in pain and demoralization. To date, PAT has not been studied among people living with both demoralization and chronic pain nor has it been studied as part of routine multidisciplinary outpatient palliative care.

**Objectives:** We conducted an open-label pilot study assessing the safety, feasibility, and acceptability of PAT delivered with multidisciplinary palliative care support in cancer patients across the illness trajectory living with demoralization and chronic pain.

**Methods:** Participants received a single 25 mg oral dose of psilocybin with preparation, monitoring, and integration provided by a mental health clinician and spiritual health clinician, alongside multidisciplinary palliative care support. Outcomes included safety, feasibility, acceptability, and exploratory self-report measures assessing for demoralization and pain intensity pre- and post-dosing.

**Results:** Eleven participants were enrolled, ten of whom received psilocybin. The intervention was safe and feasible, with no serious adverse events and complete study visit retention among dosed participants. All 10 dosed participants reported the intervention as highly acceptable. Among dosed participants, 70% rated the experience among the five most meaningful and educational of their lives, and 60% among their five most spiritually significant experiences. By study endpoint, 90% no longer met criteria for clinically-significant demoralization syndrome and had pain scores below the trial enrollment threshold.

**Conclusions:** PAT delivered with multidisciplinary palliative care support was safe, feasible, and acceptable in demoralized cancer patients with chronic pain.

**Key Message:** Psilocybin-assisted therapy delivered within multidisciplinary outpatient palliative care was safe, feasible, and acceptable among demoralized cancer patients with chronic pain.

## Introduction

Demoralization syndrome is a clinical condition and form of existential distress characterized by hopelessness, helplessness, and a loss of meaning.^1^ It is common among cancer patients at all ages and stages of disease (24-36% prevalence; >3 million adults in the US).^2,3^ Demoralization may result in a perceived failure to adaptively cope may result in a sense of failure, isolation, and suicidal thinking.^4^ In contrast to major depressive disorder, demoralization is characterized by “subjective incompetence,” which is the self-perceived incapacity to express feelings or perform tasks when facing a stressor, leading a person to feel trapped in their predicament.^5^ Importantly, demoralization is more predictive of suicidal ideation than major depressive disorder alone, signifying a serious and under-recognized health condition in cancer populations.^4,6,7^ To date, there are no evidence-based pharmacologic interventions for demoralization syndrome as a primary target of treatment.

Chronic pain (pain > 3 months) is also highly prevalent in cancer populations, impacting one in three cancer patients (>5 million adults) and is associated with poor quality-of-life, treatment non-adherence, and elevated healthcare costs.^8^ Chronic pain is associated not only with demoralization in patients receiving early palliative care, but also pain-related helplessness, death anxiety, and PTSD symptoms.^9^ Given that chronic pain and demoralization are often comorbid in cancer and palliative care populations, the study of transdiagnostic interventions (i.e., therapies that target multiple constructs at once) is attractive in palliative care to reduce polypharmacy and provide polysymptomatic relief to vulnerable populations living with both physical, emotional, and existential pain.^9,10^

Psychedelic-assisted therapy has gained renewed interest over the past two decades for its transdiagnostic potential to treat pain and numerous mental health constructs, including depression, anxiety and PTSD.^11–16^ In particular, psilocybin-assisted therapy (PAT), the combination of psilocybin (an agonist of the 5-HT2A receptor) with therapeutic approaches, has seen a groundswell in serious illness research over the past decade.^17–19^ Numerous pilot studies and randomized controlled trials (RCTs) have shown preliminary efficacy in treating symptoms of anxiety and depression,^20–23^ with one larger RCT underway in cancer populations (NCT05398484). Furthermore, PAT for cancer pain has gained interest with one ongoing open-label study for opioid-refractory pain in patients with advanced-cancer (NCT06001749). With two phase III trials studying psilocybin for depression now complete and pending publication, psilocybin may soon receive marketing approval from the US Food and Drug Administration for depression, potentially allowing for off-label prescribing of PAT for palliative care populations with demoralization and/or chronic pain.

Despite increasing interest in PAT among palliative care programs, PAT has not been studied within a multidisciplinary outpatient palliative care model that resembles real-life gold standard palliative care multidisciplinary support that provides physical, emotional, social, and spiritual care through integrated support from physicians, nurses, social workers, and healthcare chaplains. Furthermore, though spiritual, existential, religious, and theological (SERT) experiences are common in PAT and function as potential mediators of improvements in anxiety, depression, and distress related to serious illness, systems to deliver SERT-responsive care have not yet been incorporated to psychedelic-palliative care trials.^24^ PAT has also not been studied in cancer populations with comorbid chronic pain and demoralization across the illness trajectory.

Currently, there is no evidence-based model of care for integrating PAT into routine outpatient palliative care. In response to these gaps, we studied the safety, feasibility, and acceptability of PAT for demoralized cancer patients with chronic pain coupled with outpatient multidisciplinary palliative care support in which healthcare chaplains were included as core members of the PAT facilitation team.

## Methods

This study was approved by the Emory Institutional Review Board and was registered on Clinicaltrials.gov (NCT05506982). Participants were recruited from November 1, 2022 to January 9, 2024. The study was monitored by the Winship Cancer Institute Data Safety and Monitoring Board.

### Recruitment

Participants were primarily recruited from Emory’s Winship Cancer Institute and outpatient palliative care program. Additional participants were recruited from online advertisement and word of mouth. Potential participants were screened over the phone prior to presenting for a full screening visit. While not required for enrollment, participants were asked to elect a caregiver to assess the feasibility of recruiting and maintaining caregivers and to study the impact of the intervention on caregiver strain and global psychosocial functioning.

### Eligibility

Participants needed to have received a diagnosis of solid or liquid cancer made at least 1 year prior to screening, at any stage in cancer survivorship (specifically, active cancer treatment or no cancer treatment either in clinical remission or with advanced disease), prognosis of greater than six months as determined by their primary oncologist, moderate-to-severe demoralization (score of ≥ 10 on the Demoralization Scale-II, chronic pain (pain lasting > 3 months per patient report and score of ≥ 5 for average pain level on Brief Pain Inventory), age ≥ 26 years old and ≤ 85 years old, and availability of a trusted person into whose care the participant could be released following the drug administration session. *See Appendix for full inclusion/exclusion criteria*.

### Procedures

#### Study Design

The study consisted of three phases (Figure 1). In Phase 1, participants were screened to determine eligibility and to taper off any prohibited medications (full exclusion criteria in Appendix). The study PI and study psychiatrist worked with the participant’s outpatient palliative care clinicians to support medication tapering. Participants on prescribed opioid, benzodiazepine, or cannabinoid regimens were maintained on their regimens with no required taper to avoid undue burden given lack of clear interaction with PAT experience, though patients were able to elect to be tapered off these medications over the course of the trial. Participants were allowed up to four weeks to taper off prohibited medications. Phase 2 began with the baseline visit (day 0), followed by three preparatory sessions with facilitators (i.e., the mental health clinician and healthcare chaplain), psilocybin dosing (day 14), and three integration sessions ending six weeks after baseline (day 42, primary endpoint). Participants also saw an outpatient-based palliative care social worker and either an outpatient-based palliative care physician or advance practice provider twice before and twice after dosing.

**Figure 1.**
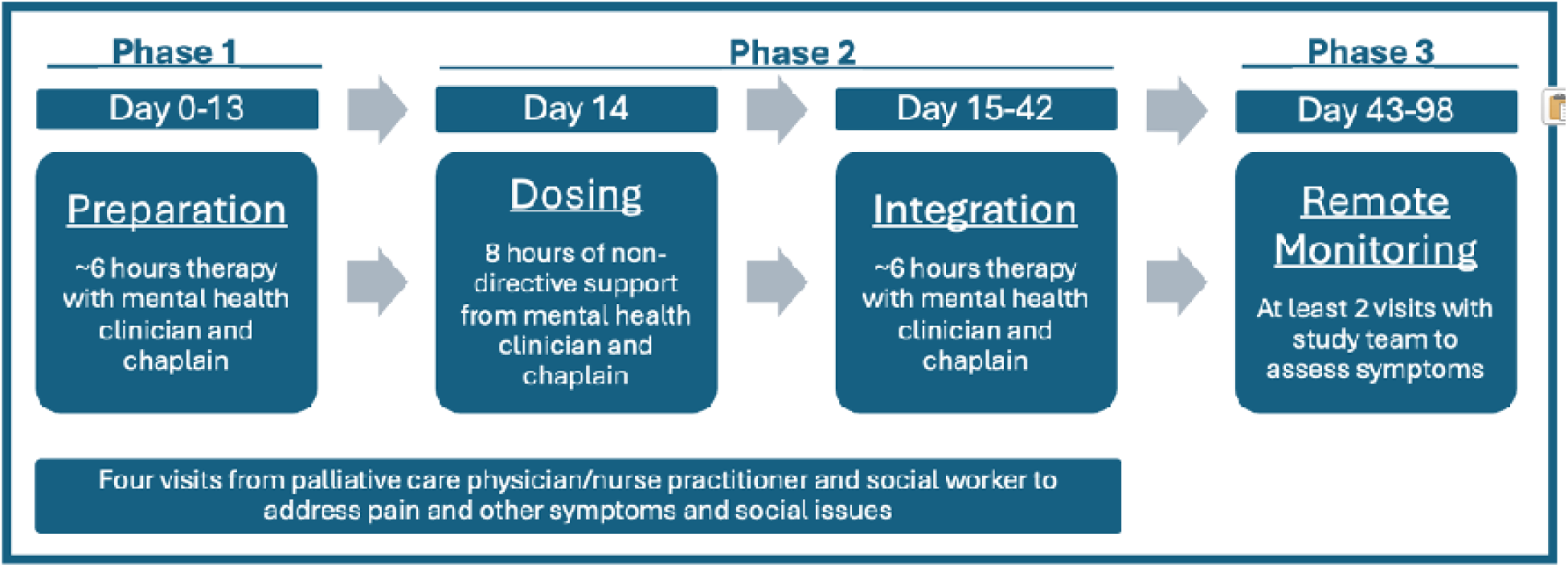
Study Design with three phases involving preparation, dosing, integration, and monitoring coupled with multidisciplinary palliative care visits.

#### PAT Facilitation Model – “Spiritually-Informed Triad Model”

Preparation and integration sessions were ∼2 hours in duration and palliative care visits were ∼1 hour in duration. Phase 3 consisted of two remote assessments collected at day 56 and day 98, to assess the feasibility of capturing adverse events and durability of the intervention on exploratory outcomes. Optional follow up with the palliative care multidisciplinary team were possible, as deemed necessary by the participant and/or study clinicians. Participants were supported by a clinical dyad of a trained mental health specialist and a certified spiritual health clinician (SHC), also known as a board-certified healthcare chaplain, who supported participants during preparation, dosing, and integration. Procedures were followed according to the Usona Institute Manual for Clinical Facilitators.^25^ In preparation sessions, the facilitators worked to establish therapeutic rapport, provide preparatory psychoeducation, explored the participant’s values and intentions for PAT, and complete a brief life review to address salient aspects of the participant’s life including their illness experience that might arise during dosing. During the dosing session, facilitators provided supportive, non-directive support. In integration sessions, facilitators worked with participants to translate insights from dosing to promote cognitive, affective, and behavioral changes consistent with the participant’s values. This pluralistic, non-imposing, and respectful approach prioritized patient autonomy and empowerment while exploring the participant’s beliefs and worldviews.^24,26,27^

SHCs did not explicitly address spirituality; however, if participants introduced spiritual, existential, religious, or theological concerns, SHCs would inquire about the significance and meaning of these concerns using a nonjudgmental stance without directiveness or imposition. This approach practiced by SHCs is consistent with the values of Compassion Centered Spiritual Health (CCSH), an evidence-based model of providing spiritual assessment and care practiced by SHCs that promotes access to compassion and cultivating attunement in both care seekers (the participant) and care responders (the facilitators), all while trusting the participant to access personal resources for healing.^28–30^ Of note, although CCSH is a manualized approach to spiritually-informed healthcare, the facilitation model in the present study used a supportive, non-directive approach recommended for psychedelic therapy and formalized in the Usona Manual.(see Appendix for detailed information SHC competencies and the Triad Model.)

#### Outpatient Multidisciplinary Palliative Care – Added Layer of Care

Participants also received an additional layer of medical, psychosocial, and spiritual support in the form of a multidisciplinary palliative care team of physicians and licensed clinical social workers (LCSWs) experienced in practicing outpatient palliative care.^31^ Physicians and advanced practice providers (APP) addressed primarily medical needs and any other issues that were not captured by the facilitator dyad. The LCSWs addressed any social issues. These clinicians documented their visits and informed the PI of any recommendations to support the participant, who then informed the facilitator dyad. The rationale for including the multidisciplinary palliative care team as an extra layer of care to support the core therapeutic triad was to bridge the study participant back to outpatient palliative care following the study’s completion, as well as to demonstrate a model of psychedelic multidisciplinary palliative care that may be implementable in real-life outpatient palliative care programs.

#### Setting

Preparation, dosing, and integration took place at Emory University in a comfortable living room-like environment with a bed for the participant and two chairs for the facilitator dyad. During dosing, participants were instructed to wear eyeshades to facilitate looking inward and to minimize outside visual distractions, though they could remove the eyeshades. A curated playlist was specifically made for this study to match the arc of a typical high-dose psilocybin experience in which more music of greater intensity was highlighted during the first three hours of the session and progressively lightened over the course of the eight-hour playlist. Fresh flowers were provided for each participant, in keeping with traditions from twentieth-century psychedelic trials and to add natural beauty to the dosing space. Participants also requested snacks of their choosing in advance of dosing day that were provided towards the end of the dosing session.

#### Study Drug

Participants were dosed with 25mg pharmaceutical grade synthetic psilocybin in gel capsules synthesized by the Usona Institute (Madison, WI, USA). In prior trials using high-dose oral psilocybin in people with serious illness, study participants were dosed either by weight (0.3-0.36mg/kg or 22 or 30 mg/70 kg) or 25mg dose with no serious adverse effects attributable to psilocybin.^32–34^ High-dose psilocybin was used instead of a lower-dose given findings of persisting benefit from 1-2 doses of high-dose psilocybin for symptoms of anxiety and depression in prior trials.^25,35,36^

#### Primary Outcomes

##### Adverse Events

Adverse events were recorded using a variety of assessments. At each study visit, suicidality was assessed using the Columbia Suicide Severity Rating Scale (C-SSRS).^37^ Medical or psychiatric treatment-related adverse events were collected using the National Cancer Institute-Common Terminology Criteria for Adverse Events (CTCAE) at all study visits and all points of contact with the participant. On dosing day, facilitators collected vital signs at one-hour intervals throughout the session. The facilitators also monitored for adverse events (AEs) as defined by the CTCAE throughout the session. Attribution of the AE’s relatedness to psilocybin was determined by the study PI and informed by its proximity to the acute effects of dosing.

##### Feasibility and Acceptability

Feasibility was determined by participant retention in the study and number of missed visits. The pre-specified threshold for successful participant retention in the study and number of missed events was 80%. Acceptability of the intervention was assessed using 1) a single-item question asking participants’ overall satisfaction of the intervention (“Overall, I am satisfied with this intervention,” scored from 1=strongly disagree to 5=strongly agree) and 2) a Retrospective Questionnaire used in prior PAT studies in which participants provided discrete and free-text responses to memorable, meaningful, educational, and/or spiritual experiences attributable to the intervention completed at Day 14 (end of dosing day), Day 42, and Day 98 (See Appendix for specific questions).^35^ The predetermined threshold for successful acceptability was that 60% of participants would find the intervention satisfactory.

#### Exploratory Outcomes

The exploratory objectives of this study were to evaluate for changes in demoralization, pain-related outcomes (intensity and interference, pain catastrophizing), anxiety, depression, and multidimensional flourishing in multiple domains including happiness and life satisfaction, physical and mental health, meaning and purpose, character and virtue, and close social relationships. We also collected potential predictors of outcomes including mystical-type experience and challenging experiences related to psychedelic use. Caregiver psychosocial functioning and stress were also assessed and will be reported in another paper. As chronic pain and demoralization were key inclusion criteria, these constructs, as well as depression and pain catastrophizing which are highly comorbid with chronic pain and demoralization, were the exploratory outcomes of primary interest. We characterized participants’ pain phenotypes using the International Association for the Study of Pain (IASP) guidelines for nociplastic pain in which the study physician used clinical exam and history to determine if participants and nociceptive, neuropathic, nociplastic, or mixed pain.^38–40^ These constructs, as well as overall life satisfaction, were studied using the following measures:

#### Brief Pain Inventory (BPI)

The BPI assesses the severity of pain and its impact on function (Chronbach’s alpha for pain intensity reliability range 0.77-0.85). Pain items include worst and least pain in the last 24 hours, average pain level, and current pain level.^41^

#### Demoralization Scale-II (DS-II)

The DS-II is an abbreviated, 16-item version of the well-validated original 24-item Demoralization Scale which is the most widely used self-report measure of demoralization in palliative care patients. The DS-II has shown both good internal and external validity in palliative care patients (Cronbach’s alpha range 0.81-0.93).^42^

#### Hospital Anxiety and Depression Scale (HADS)

The HADS is a 14-item self-report scale and reliable instrument for detecting states of depression and anxiety in outpatient settings, each yielding a maximum score of 21 with higher scores representing more severe anxiety or depression. Cronbach’s alpha ranges from 0.68-0.93 for the anxiety subscale and 0.67-0.90 for the depression subscale.^43^

#### Pain Catastrophizing Scale (PCS)

The PCS is a self-report measure that assesses maladaptive pain consisting of 13 items scored from 0 to 4. Possible scores range from 0 to 52. Higher scores indicate that the individual tends to catastrophize to a greater degree (Cronbach’s alpha ∼0.92). The measure consists of three validated subscales: rumination, magnification, and helplessness.^44^

#### Harvard Flourishing Measure (HFM)

The HFM is a self-report measure based around five central domains: happiness and life satisfaction, mental and physical health, meaning and purpose, character and virtue, and close social relationships. Life satisfaction was assessed by the single-item metric from the HFM used to assess this construct: “Overall, how satisfied are you with life as a whole these days?” and is scored from 0 to 10 (highest satisfaction).^45^

We explored experiences related to the acute psilocybin experience with the following measures administered at the end of dosing day (Day 14):

#### Mystical Experience Questionnaire (MEQ)

The MEQ is 30-item 6-point (0-5) self-report measure of mystical-type experiences developed within the context of psychedelic research that captures domains related to mystical experience (e.g., feelings of experiencing eternity or infinity, sense of reverence, experience of oneness, experiences of the sacred and holy), positive mood, transcendence of space and time, and ineffability of experience (i.e., sense that the experience cannot be described adequately in words). The maximum score is 150.^46^

#### Challenging Experience Questionnaire

The CEQ provides a phenomenological profile of challenging aspects of experiences with psilocybin such as grief, fear, death, insanity, isolation, physical distress, and paranoia. Subscale scores have been associated with difficulty, meaningfulness, spiritual significance, and change in well-being attributed to the challenging experiences.^47^

#### Qualitative Interviews

Postintervention interviews using the Enhanced-Critical Incident Technique were conducted with study participants, their caregivers, and study facilitators to understand components of the intervention that were salient to outcomes (either perceived positive or negative) as well as wish-list items that were desired but not part of the protocol. Findings from our qualitative analysis have been published.^48^ In the Retrospective Questionnaire, participants were asked to describe the most memorable, meaningful, educational, and/or spiritually significant experiences related to their dosing session. We synthesized illustrative narratives of the dosing day experience for this manuscript.

#### Sample Size and Data Analysis

We chose a sample size of 10 dosed participants based on prior PAT feasibility studies and practical considerations, with the primary goal of the study being 10 of 10 patients completing the dosing regimen and follow-up. Thus, statistical inference has not been performed as the study was not powered to test a particular hypothesis or provide specific widths of confidence intervals. Rather, descriptive statistics were used for the primary outcomes including rates of recruitment, retention, baseline clinical characteristics, and adverse events. Additionally, continuous efficacy outcomes are summarized by time-point using medians and interquartile ranges at baseline, study endpoint (day 42), and post-study follow-up (day 98). Cohen’s d statistic to capture the effect size. Analyses were performed using SAS software (version 9.4; SAS Institute Inc., Cary NC).

## Results

Consistent with target recruitment in pilot feasibility studies, our recruitment target was 10 individuals. Of the 30 individuals who expressed interest in the study, 26 were prescreened, and 16 were eligible for screening. Of the 15 who were eligible, one did not wish to enroll upon learning more about the study, three did not qualify for the study after consenting (two with QTc > 450ms and one with bipolar II diagnosis), and 11 moved forward with baseline preparatory visits. One participant withdrew from the study between the second and third preparatory visits as his goals became focused on pursuing PAT as an antineoplastic intervention which was not consistent with the goals of the study. Of the 11 enrolled in the study, 10 were dosed and completed the study up to day 98 follow up (Figure 2). Of the 10 patients who completed the protocol, seven were female and nine were white (Table 1). All but one had a history of major depressive disorder (six current and three in remission) and two each had panic disorder, generalized anxiety disorder, and PTSD. Six participants had advanced cancer of whom four were on active cancer-directed therapy. Two had graft vs. host disease following bone marrow transplant. Seven of 10 participants elected to taper off contraindicated medications prior to baseline, the most common being duloxetine (four of 10). Participants had a mix of pain phenotypes with the most common being a mixed phenotype, with three having comorbid nociceptive and neuropathic pain and two having comorbid nociceptive, neuropathic, and nociplastic pain.

**Figure 2.**
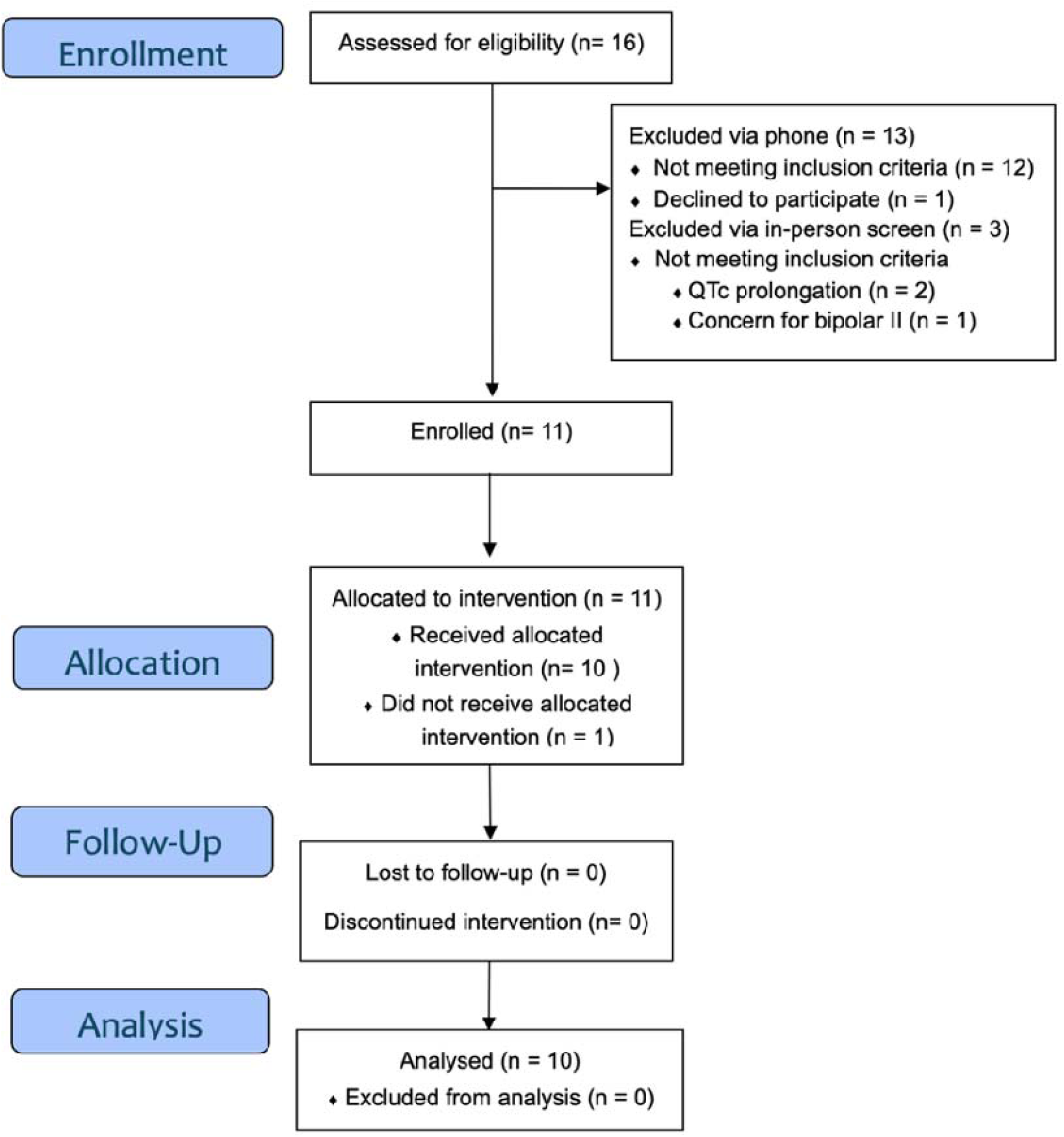
CONSORT Flow Diagram.

**Figure 3.**
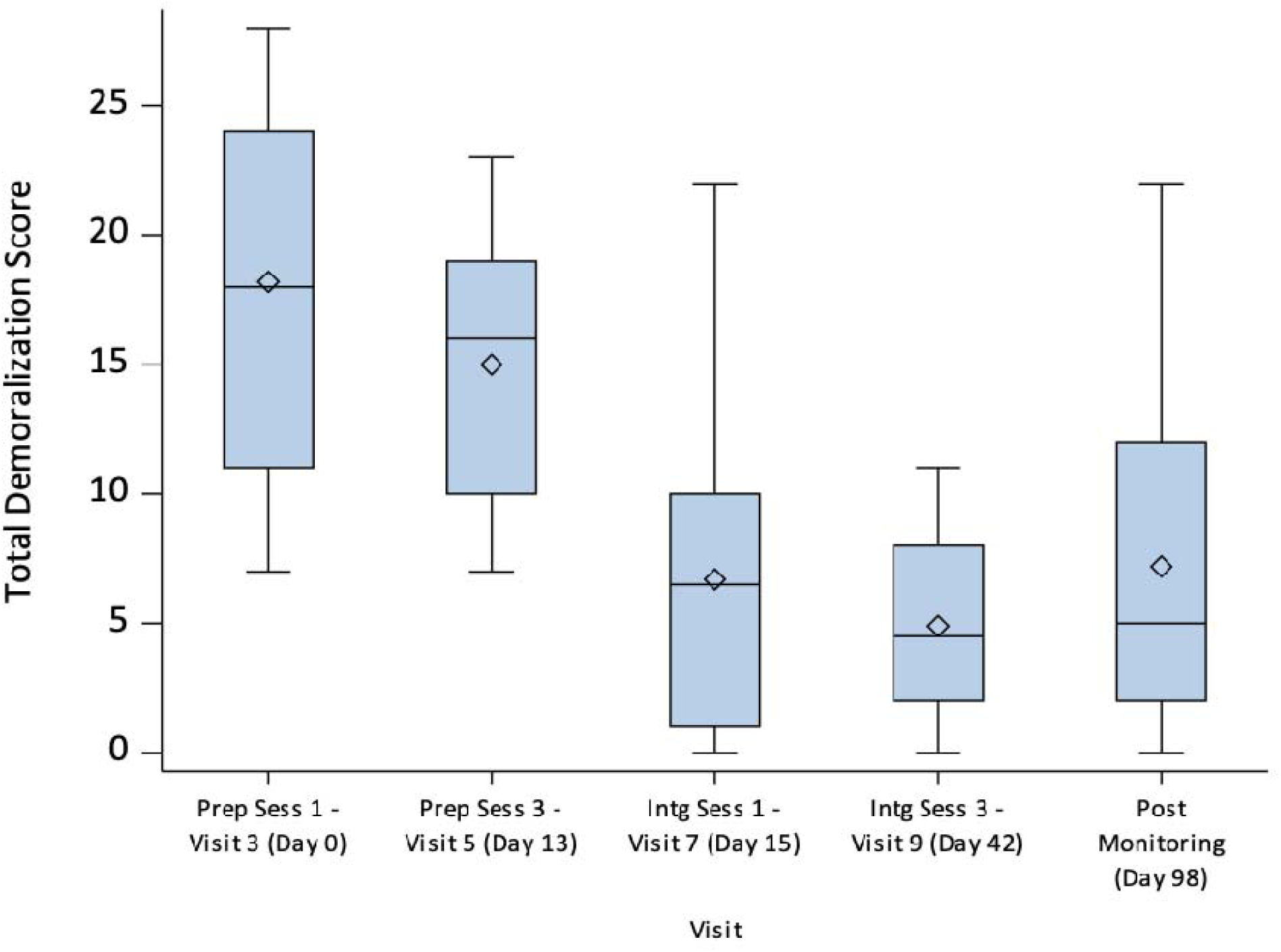
Demoralization Scores at baseline and Day 13 (day pre-dosing), 15 (day post-dosing), 42 and 98 presented as box plots with interquartile range (IQR). Dashed line represents the median and diamond represents the mean.

**Figure 4.**
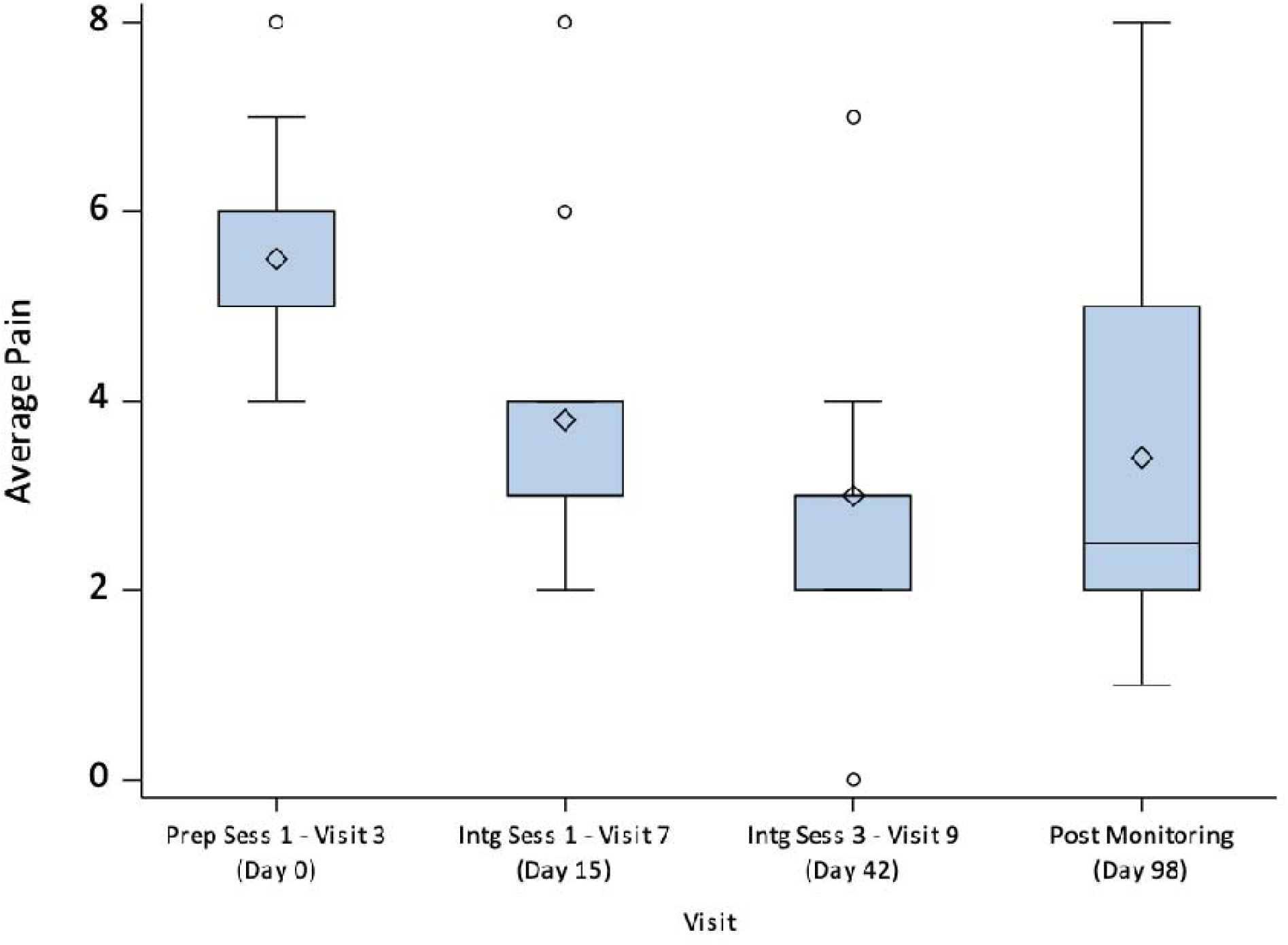
Average Pain Scores at baseline, Day 15 (day post-dosing), 42 and 98 presented as box plots with interquartile range (IQR). Dashed line represents the median and diamond represents the mean.

**Table 1.** Demographic and Diagnostic Characteristics.

| **Demographic Characteristics** |  |
| --- | --- |
| Variable | Descriptive statistics |
| Age: Mean (SD), range | 53.90 (12.84), 29-69 |
| Gender | 7 female, 3 male |
| Race | 9 White, 1 multi-racial |
| Ethnicity | 10 non-Hispanic |
| Highest Educational Level | Some College (1), Bachelor's Degree (4), Master's Degree (3), Doctoral Degree (2) |
| Household Income | >100K (6), 80-100K (2), 60-80K (1), <20K (1) |
| **Diagnostic Characteristics** |  |
| Time since diagnosis in years: Mean (SD), range | 8.90 (6.77), 3-17 |
| Cancer type and related syndromes | Acute myeloid leukemia complicated by graft vs. host disease, breast cancer (3), Ewing's sarcoma, multiple myeloma, myelodysplastic syndrome complicated by graft vs. host disease, ovarian cancer (2), peritoneal cancer |
| Disease stage | Advanced, active treatment (4) |
|  | Advanced, no evidence of disease (2) |
|  | Non-advanced, active treatment (2) |
|  | Non-advanced, surveillance (2) |
| Comorbid psychopathology | Major Depressive Disorder (current (1), past (3), current and past (recurrent) (5)) |
|  | Panic disorder (2) |
|  | Generalized Anxiety Disorder (2) |
|  | Post-Traumatic Stress Disorder (PTSD) (2) |
|  | Agoraphobia (1) |
|  | Social anxiety disorder (1) |
|  | Social phobia (1) |
| Medication tapers prior to baseline visit | Duloxetine (4), Bupropion (1), Bupropion-Dextromethorphan (1), Escitalopram (1), buprenorphine (1)*** |
| Pain phenotype** | Nociceptive only (3), Neuropathic only (0), Nociplastic only (1), Mixed phenotype (6 total: nociceptive and neuropathic (3), nociceptive and nociplastic (1), nociceptive, neuropathic, and nociplastic (2) |

SD = standard deviation; \*\*based on clinician evaluation; \*\*\*tapered by participant request

### Safety

There were no serious adverse events (AEs) related to psilocybin or the intervention (Table 2). There were no changes in suicidal ideation as detected by the C-SSRS. All participants reported at least one AE during the dosing session, the most common being nausea and headache. Blood pressure and heart rate were monitored hourly over eight hours during the dosing session. There was no clinically meaningful rise in either systolic or diastolic blood pressure over the course of the dosing session. The peak systolic blood pressure was 177 mmHg and peak diastolic blood pressure was 105mmHg. The mean peak systolic blood pressure was 143 mmHg (SD 22.63) and peak mean diastolic blood pressure was 87 mmHg (SD 6.81). The peak heart rate was 96 beats per minute with peak mean of 76 beats per minute (SD 21.21). See Appendix for full list of AE’s and hourly data of heart rate and blood pressure.

**Table 2.** Adverse Events.

| Table 2. Adverse Events |  |  |  |  |  |  |
| --- | --- | --- | --- | --- | --- | --- |
| Event | n | % | Maximum Severity |  |  | Relatedness to Psilocybin* |
|  |  |  | Mild | Moderate | Severe |  |
| Nausea | 6 |  | 3 | 3 | 0 | 1 (4), 2 (2) |
| Headache | 5 |  | 3 | 2 | 0 | 1 (3), 2 (1), 3 (1) |
| Irritability | 2 |  | 1 | 1 | 0 | 1 (1), 3 (1) |
| Tinnitus | 2 |  | 2 | 0 | 0 | 2 (1), 3 (1) |
| Vomiting | 2 |  | 2 | 0 | 0 | 2 (2) |
| Hypertension | 2 |  | 1 | 1 | 0 | 3 (2) |
| Urinary Tract Infection | 2 |  | 0 | 2 | 0 | 3 (2) |
| Paresthesia | 2 |  | 2 | 0 | 0 | 1 (1), 2 (1) |
| Derealization | 1 |  | 1 | 0 | 0 | 1 (1) |
| Geometric Visuals | 1 |  | 1 | 0 | 0 | 2 (1) |
| Dizziness | 1 |  | 1 | 0 | 0 | 2 (1) |
| Insomnia | 1 |  | 0 | 1 | 0 | 2 (1) |
| Lightheadedness | 1 |  | 1 | 0 | 0 | 2 (1) |
| Gait Unsteadiness | 1 |  | 1 | 0 | 0 | 2 (1) |
| Breast pain | 1 |  | 1 | 0 | 0 | 2 (1) |
| Neck Pain | 1 |  | 1 | 0 | 0 | 2 (1) |

\* 1= related, 2=possibly related, 3=unrelated.

### Feasibility and Acceptability

The study met feasibility and acceptability milestones. All participants who were dosed with psilocybin completed study visits and ten of 11 participants who began preparation for dosing (baseline visit) completed the trial. All primary assessments were completed for all participants who were dosed. Among exploratory assessments, there were only four assessments that had missing data. All participants who were dosed rated the intervention as highly acceptable (100%). Seven of 10 rated the intervention as among the top five most meaningful and educational experiences of their lives, with one reporting it as the single most meaningful experience of their life and two reporting it as the single most educational experience of their life. Six of 10 rated the intervention as among the top five most spiritually significant experiences of their lives, with one reporting it as the single most spiritually significant experience of their life.

### Exploratory Outcomes

#### Demoralization

Large reductions in demoralization were observed from baseline (Day 0) to study endpoint (Day 42) (median change from baseline -14 points (IQR -19, -8), *Cohen’s d* -0.94) and up to Day 98 follow-up (median change - 11.5 points (IQR -18, -6), *Cohen’s d* -1.49). Nine of 10 of participants no longer met criteria for clinically-significant demoralization syndrome at Day 42 (DS-II score < 10) and six did not meet criteria at Day 98. Participants also had pronounced decreases in depression scores (Table 3).

**Table 3.** Exploratory Outcomes.

| Table 3. Exploratory Outcomes |  |  |  |  |  |
| --- | --- | --- | --- | --- | --- |
|  |  | Raw Score |  | Change from Baseline |  |
| Outcome | Time | N | Median (IQR) | N | Median (IQR) |
| **DS-II** |  |  |  |  |  |
|  | D0 | 10 | 18 (11, 24) | 10 | 0 (0, 0) |
|  | D13 | 10 | 16 (10, 19) | 10 | -2.5 (-7, -1) |
|  | D15 | 10 | 6.5 (1, 10) | 10 | -10.5 (-15, -6) |
|  | D42 | 10 | 4.5 (2, 8) | 10 | -14 (-19, -8) |
|  | D98 | 10 | 5 (2, 12) | 10 | -11.5 (-18, -6) |
| **HADS – Depression** |  |  |  |  |  |
|  | D0 | 10 | 12 (9, 15) | 10 | 0 (0, 0) |
|  | D13 | 10 | 10.5 (8, 13) | 10 | 0 (-3, 2) |
|  | D15 | 10 | 6.5 (5, 9) | 10 | -3.5 (-4, -2) |
|  | D42 | 10 | 4.5 (2, 10) | 10 | -5 (-10, -1) |
|  | D98 | 10 | 4.5 (1, 10) | 10 | -3.5 (-13, -1) |
| **HADS – Anxiety** |  |  |  |  |  |
|  | D0 | 9 | 10, (5, 15) | 9 | 0 (0, 0) |
|  | D13 | 10 | 10.5 (4, 13) | 9 | -2 (-2, 1) |
|  | D15 | 10 | 4 (4, 8) | 9 | -4 (-5, -1) |
|  | D42 | 10 | 4 (1, 7) | 9 | -3 (-9, -2) |
|  | D98 | 10 | 5 (4, 8) | 9 | -2 (-8, 1) |
| **BPI – Average Pain Severity** |  |  |  |  |  |
|  | D0 | 10 | 5 (5, 6) | 10 | 0 (0, 0) |
|  | D15 | 10 | 3 (3, 4) | 10 | -1 (-3, -1) |
|  | D42 | 10 | 3 (2, 3) | 10 | -2.5 (-3, -1) |
|  | D98 | 10 | 2.5 (2, 5) | 10 | -2 (-4, 0) |
| **PCS** |  |  |  |  |  |
|  | D0 | 10 | 25.5 (16, 35) | 10 | 0 (0, 0) |
|  | D13 | 10 | 29.5 (21, 33) | 10 | 1 (-3, 3) |
|  | D15 | 10 | 14 (1, 22) | 10 | -11.5 (-30, 0) |
|  | D42 | 10 | 4 (1, 17) | 10 | -15 (-31, -12) |
|  | D98 | 10 | 7.5 (2, 19) | 10 | -13.5 (-32, 1) |
| **HFM – Overall Life Satisfaction** |  |  |  |  |  |
|  | D0 | 10 | 2.5 (1, 5) | 10 | 0 (0, 0) |
|  | D42 | 10 | 8 (4, 8) | 10 | 3.5 (2, 6) |
|  | D98 | 10 | 7 (3, 9) | 10 | 2.5 (1, 6) |

D0=baseline, D13=day prior to dosing, D15=day after dosing, D42=study endpoint, D98=last follow up. IQR=interquartile range

#### Pain

Pain intensity scores declined from baseline (Day 0) to study endpoint (Day 42) (median change -2.5 points (IQR -3, -1), *Cohen’s d* -1.46) and up to Day 98 follow-up (median change -2 points (IQR -4, 0), *Cohen’s d* - 1.13). Nine of 10 of participants had pain scores lower than cutoff for trial enrollment (BPI average pain intensity < 5) at Day 42, and seven of 10 had scores lower than cutoff at Day 98. Of note, the same nine participants who no longer met criteria for clinically-significant demoralization syndrome also had BPI average pain intensity scores < 5 at Day 42. Participants also had pronounced decreases in the pain catastrophizing total score as well as pain magnification, rumination, and helplessness subscales (Table 3 and Appendix).

#### Life Satisfaction

The overall life satisfaction score, based on the single item from the HFM, increased from a median of 2.5 (IQR 1,5) at baseline to 8 (IQR 4,8) at study endpoint, and 7 (IQR 3,9) at Day 98 follow-up (Table 3 and Appendix).

#### Dosing Day Experiences

The median total score of the Mystical Experience Questionnaire (MEQ30) was 123 (IQR 91, 139). Median scores in the mystical, positive mood, transcendence of space/time, and ineffability were similarly high (Table 4). The highest median subscales of the Challenging Experience Questionnaire were grief followed by physical distress and fear.

**Table 4.**
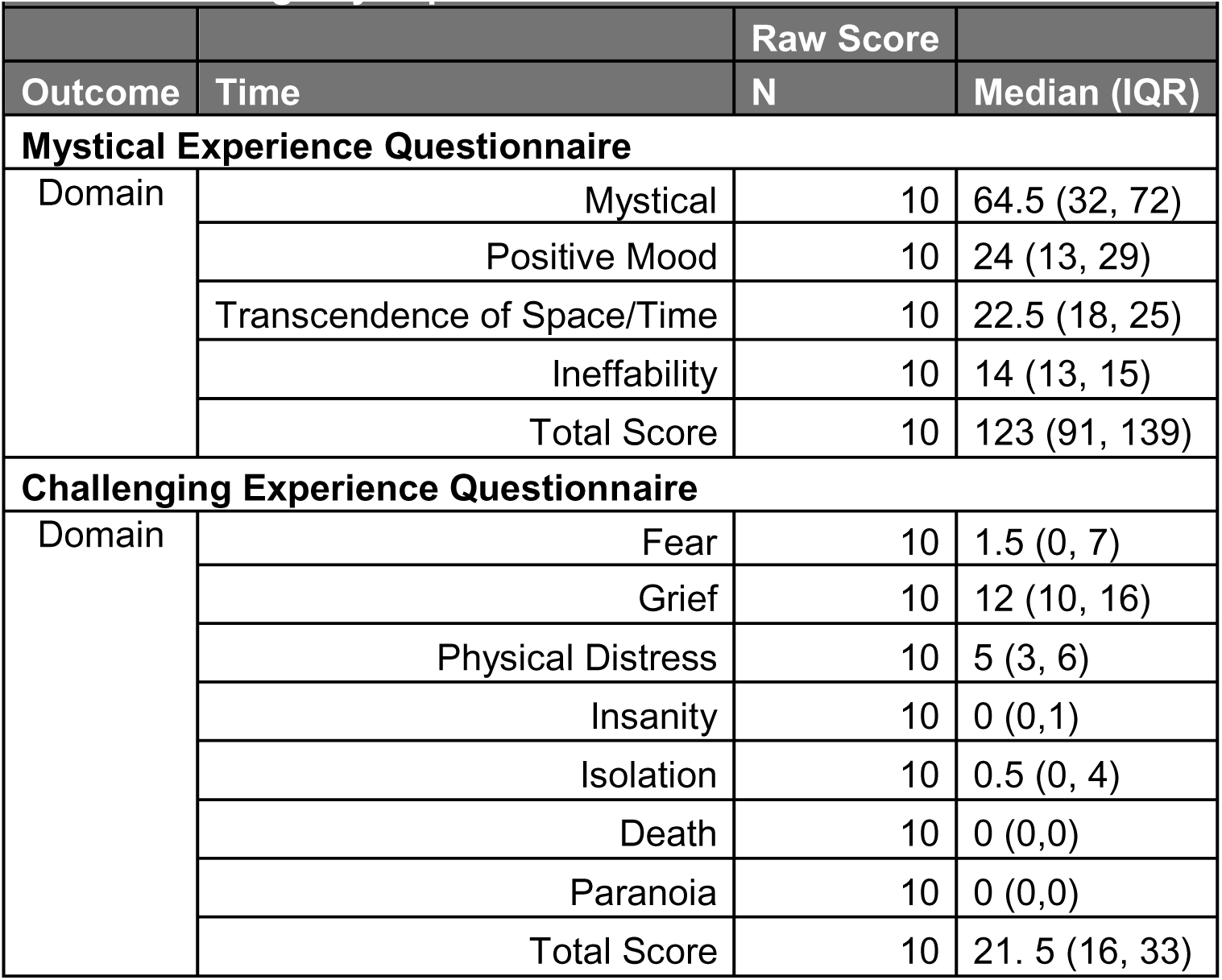
Dosing Day Experiences.

### Medication Taper Follow up

Of the seven participants who tapered prohibited medications for the study, five restarted their medications by Day 98. Two did not restart the duloxetine. The participant who tapered off buprenorphine remained off it at Day 98.

## Discussion

This pilot study demonstrated strong safety, feasibility, and acceptability of delivering psychedelic-assisted therapy (PAT) alongside multidisciplinary palliative care support among people living with cancer with clinically-significant demoralization and chronic pain across the illness trajectory. It is also the first PAT study to incorporate spiritual health clinicians as facilitators providing preparation, dosing, and integration support to all participants.^24,26,27^

This study is unique in enrolling people living with cancer and chronic pain comorbid with demoralization. Both pain and demoralization are highly prevalent and frequently co-occur among people living with serious illness receiving outpatient palliative care.^9,10^ Within this small cohort, most, but not all, participants experienced meaningful and durable reductions in demoralization and pain severity. As chronic pain in cancer populations is complex and multidimensional and often only partially responsive to pharmacological analgesics, PAT may be of great potential to simultaneously reduce psychological distress and modulate the subjective experience and impact of chronic pain.^13,49^ As a single, multicomponent intervention, PAT may represent a paradigmatic change to alleviating distress in palliative care rather than pursuing conventional symptom-specific treatments for comorbid distress.

These findings complement a growing body of research examining PAT for demoralization among people living with serious illness. A recent pilot study demonstrated the feasibility of integrating PAT within interdisciplinary hospice care for patients with a life expectancy of less than six months and clinically significant demoralization.^20^ Similarly, another study demonstrated the feasibility of PAT among AIDS survivors with clinically significant demoralization.^22^ Collectively, these open-label studies support the safety and feasibility of PAT for seriously ill populations experiencing demoralization and suggest that demoralization may be an important therapeutic target for PAT interventions in palliative care.^50^ Importantly, a novel contribution of this study is inclusion of a SHC along with a mental health clinician within this PAT model. Our findings suggest the important role of including SHCs within psychedelic care. These findings warrant further evaluation in larger randomized trials designed to assess efficacy.^26,27^

In our cohort, six participants also met DSM-5 criteria for major depressive disorder, which, although conceptually distinct from demoralization syndrome, frequently co-occurs in seriously ill populations.^4,7,51^ Future studies should investigate whether PAT differentially affects outcomes in patients with demoralization syndrome compared to those with major depressive disorder alone. These findings also support the continued use of clinician-administered psychiatric assessments alongside patient-reported outcomes in palliative care trials to identify comorbid psychiatric diagnoses and provide more objective assessment of psychiatric constructs (vs. patient-reported outcomes only) in medication and behavioral intervention serious illness research.^52,53^

At study endpoint, nine of 10 dosed patients no longer met criteria for clinically-significant demoralization syndrome and had pain and demoralization scores lower than cutoff for trial enrollment. However, interquartile ranges and confidence intervals were wide, which are unsurprising for a pilot feasibility study. Furthermore, participants presented with a range of pain phenotypes, most commonly involving overlapping nociceptive and neuropathic pain. Future studies should investigate whether specific pain phenotypes respond differently to PAT or tailor PAT to specific phenotypes.^38,40,54^ An important component of this research agenda will be determining whether certain pain phenotypes are more strongly associated with psychological and psychospiritual distress, and whether reductions in demoralization might mediate improvements in pain, or vice versa. Importantly, the adapted Usona manual used in this pilot study did not explicitly provide any pain-specific modules or tools to help participants manage pain, and still most participants reported improvements in pain. The addition of pain-specific modules, such as pain neuroscience education, during preparation and integration, might bolster the effects of PAT on pain impact.^49^ Future studies might evaluate the additive effects of such modules on potency and durability of treatment response.

Most participants were tapered off prohibited medications prior to study participation, and most resumed these medications following dosing. It is important to explore circumstances when foregoing such tapers may be appropriate. Recent findings suggest that PAT may still produce acute alterations in consciousness and improvements in mental health symptoms among patients maintained on serotonergic medications (e.g., SSRIs and SNRIs), suggesting that tapers might not always be necessary to achieve therapeutic effects in PAT.^55,56^

This study was completed while two phase III trials investigating psilocybin for major depressive disorder and treatment-resistant depression were nearing completion. Psilocybin may soon receive FDA approval, potentially allowing clinicians to deliver PAT off-label for seriously ill patients experiencing chronic pain and/or demoralization. Several states have also legalized psilocybin and are developing clinical programs and protocols for seriously ill populations, including New Mexico.^57^ In addition, a recent Executive Order and ongoing litigation filed by palliative care clinicians seeking to reschedule psilocybin may further expand access to PAT for seriously ill patients through Right to Try pathways.^58^ This study testing the feasibility and acceptability of including SHCs is an important contribution in advancing the potential need to scale PAT in a more broad fashion in which two mental health clinicians may represent a barrier to access.^59^

This rapidly evolving therapeutic and regulatory landscape underscores the importance of conducting trials that reflect real-world applications of PAT within palliative care settings. This intervention was delivered by a multidisciplinary team of experienced palliative care clinicians with established expertise in supporting the biopsychosocial and spiritual needs of seriously ill patients. The high acceptability of this intervention, together with accumulating evidence supporting the safety, feasibility, and preliminary efficacy of PAT for depression and demoralization in cancer populations, suggests that palliative care should prioritize investigating multidisciplinary, medication-assisted interventions such as PAT integrated with palliative care.

As legislation evolves and FDA approvals potentially expand access to psychedelic therapies for people living with serious illness, a dedicated PAT research agenda within palliative care could support the development of standardized, manualized interventions targeting specific clinical outcomes, including pain, demoralization, and depression, while also informing specialized training programs for multidisciplinary palliative care teams.

### Limitations

Consistent with other pilot feasibility studies, this study is not generalizable, nor does it establish efficacy due to its small sample size, narrow geographic region (participants recruited from two southeastern outpatient palliative care clinics), and economic and racial/ethnic homogeneity of the sample. Only descriptive statistics were used without examining associations between exposures and outcomes. The exploratory outcomes of pain and demoralization, though compelling, should be interpreted as preliminary as there was no control group, blinding, randomization, or adequate statistical power for inference.

## Conclusion

This study demonstrates the feasibility and acceptability of PAT delivered by a multidisciplinary palliative care team in demoralized cancer patients with chronic pain. While safety cannot be determined by a pilot feasibility trial, there were no serious adverse events (SAE’s) attributable to psilocybin and no SAE’s during the entirety of the trial. In this small sample, most participants had robust reductions in demoralization and pain, demonstrating some strong treatment responders and nonresponders, necessitating further research to identify predictors of response as well as randomized blinded efficacy trials in more diverse populations. Facilitation support from a mental health clinician and SHC dyad was feasible and acceptable to participants. Our findings highlight the potential value of, and need for, further research examining PAT within routine outpatient early palliative care for the treatment of pain and psychospiritual distress.

## Data Availability

All data produced in the present study are available upon reasonable request to the authors

## Acknowledgements

This study was made possible through the generous support of numerous clinicians who cared for participants throughout the trial and continued to support them in clinical settings after its completion. We extend our deep gratitude to the outpatient palliative care clinicians who supported study participants, including physicians Drs. Kimberly Curseen, Lawson Marcewicz, and Nupur Dalal; nurse practitioner Courtney Cawthon; mental health specialists Shannon Jones, Dr. Mary Gresham, and Dr. Helen Vantine; social worker Stephanie Choate LCSW; and spiritual health clinicians Margaret Drew Bongiovanni, Dr. George Grant, Dr. Caroline Peacock, and Raymond Walker III. We also thank Drs. Justin Trop and Zach Burkhardt for establishing a post-trial support group for participants, which will be the subject of a future publication. We are grateful to our clinical research coordinators, Florencia Scarfone, Kaila Flemming-Peruza, and Rachel Spataro, for their invaluable contributions; Nupur Dalal and then-students Dr. Fayzan Rab and Isabelle Shub for their assistance with data entry and manuscript preparation; Dr. Malynn Utzinger of the Usona Institute for her support with training our team in the safe and ethical use of psychedelic therapies; and, finally, we thank the Mood and Anxiety Disorders Program in the Department of Psychiatry and Behavioral Sciences and Emory Spiritual Health for supporting this palliative care clinical trial.

## Funding

Research reported in this publication was supported in part by Developmental Funds from the Winship Cancer Institute of Emory University. Research reported in this publication was supported in part by the biostatistics shared resource, a core supported by the Winship Cancer Institute of Emory University under award number P30CA138292. The content is solely the responsibility of the authors and does not necessarily represent the official views of the National Institutes of Health.

## Ethics Approval

This study involved human participants and was approved by the Emory University Institutional Review Board on August 2, 2022.

## Appendix

### I. Full Inclusion and Exclusion Criteria

#### Inclusion Criteria

Participants needed to have a 1) diagnosis of solid or liquid cancer made ≥1 year at any stage in cancer survivorship (specifically, active cancer treatment or no cancer treatment either in clinical remission or with advanced disease), 2) prognosis of greater than six months as determined by their primary oncologist, 3) moderate-to-severe demoralization (score of ≥ 10 on the Demoralization Scale-II, 4) chronic pain (pain lasting > 3 months per patient report) and score of ≥ 5 for average pain level on Brief Pain Inventory, age ≥ 26 years old and ≤ 85 years old, and 5) availability of a trusted person into whose care the participant can be released following the drug administration session. Of note, we registered and advertised this study for “cancer survivors” using the definition of individuals with cancer from time of diagnosis and for the balance of life, including post-treatment survivorship and end-of-life, though we excluded participants with a life expectancy of < 6 months to allow for adequate time to complete the study procedures and because the focus of this study was on assessing the feasibility of PAT in early palliative care (not end-of-life).

#### Exclusion Criteria

Exclusion criteria included pregnancy or breastfeeding; women of childbearing potential (i.e., not permanently sterilized, not postmenopausal) who declined to use a highly effective dual contraceptive method for the duration of the study; age < 26 years old and > 85 years old; poor functional status (Eastern Cooperative Oncology Group (ECOG) score of ≥ 3); major cognitive impairment as determined by principal investigator; non-fluency in the English language; personal history of a psychotic disorder or bipolar disorder type I/II; active suicidal ideation with intent in the last 3 months (Columbia Suicide Severity Rating Scale suicidal ideation score >3) or any suicide attempt in the past year; current substance use disorder (i.e., present in last six months), excluding tobacco use disorder, of greater than mild severity as defined by Diagnostic and Statistical Manual of Mental Disorders (DSM-V); history of a seizure disorder in adulthood; active central nervous system (CNS) metastases or symptomatic CNS infection; uncontrolled hypertension (mean blood pressure exceeding 139 mmHg systolic and 89 mmHg diastolic) and heart rate exceeding 90 beats per minute at screening; clinically significant cardiovascular disease (coronary artery disease, congestive heart failure, arrhythmia, or QTc>450ms); supplemental oxygen requirement; Body Mass Index ≤18; renal insufficiency as evidenced by creatinine clearance (CrCl) <30 mL/min; considered by the principal investigator to be inappropriate for the study due to safety or to be unlikely to complete the protocol; concomitant use of drugs known to interact with psilocybin (probenecid, diclofenac); consistent use of serotonergic drugs including selective serotonin reuptake inhibitors (SSRIs), serotonin-norepinephrine reuptake inhibitors (SNRIs), or efavirenz, as well as monoamine oxidase inhibitors (MAOIs). Participants using any other antidepressant, stimulant, or antipsychotic medications on Day 0 (baseline visit) were allowed to enroll in the study if they elected to taper off the medications under medical supervision by the day prior to the baseline visit. Co-morbid psychiatric comorbidities including conditions that would exclude a potential participant were assessed by the PI and study psychiatrist using the Mini-International Neuropsychiatric Interview (MINI). The tapering period for contraindicated medications was four weeks and was managed by the PI and study psychiatrist.

### II. Facilitator Qualifications

Per FDA guidelines for psychedelic drug trials, one member of the triad was a clinician with graduate-level professional training and clinical experience in psychotherapy, licensed to practice independently. All mental health clinicians in this study had experience facilitating in PAT clinical trials. Spiritual Health Clinicians (SHCs) in this study were credentialed by the Association for Professional Chaplains (APC) as Board Certified Chaplains and/or Association for Clinical Pastoral Education (ACPE) as ACPE-Certified Educators. All had at minimum a Master of Divinity, residency in ACPE Clinical Pastoral Education and were certified as Compassion-Centered Spiritual Health (CCSH)-Registered Clinicians. All facilitators involved in this study completed additional training in PAT in July 2022 by the Usona Institute which included prerecorded videos and a live virtual two-day training on set and setting approaches to PAT, aspects of the study protocol, assessment of adverse events, a review of case studies, and supporting participants through preparation, dosing, and integration. Cultural competence and cultural safety were addressed minimally through training with the Usona manual, though core members of the study team attended workshops on implicit bias during this study. Palliative care clinicians and SHCs separately completed modules on cultural competence and safety as part of their training and employment at Emory University and Emory Healthcare. SHCs received training through Clinical Pastoral Education in socio-cultural identity, spiritual/values-based orienting systems, justice-seeking awareness of bias, intercultural and interreligious humility, and the use of cultural, religious and spiritual resources.

### III. Rationale for including a Spiritual Health Clinician as Co-Facilitator

The incorporation of a Spiritual Health Clinician (SHC) as a core member of the care team, while considered gold-standard in palliative care, is novel in psychedelic therapies. SHC have specialized knowledge and experience maintaining presence and providing guidance to seriously ill patients and their religious and spiritual needs. Qualitative research examining SHC’s unique and general contributions to PAT revealed themes of competency to work with spiritual material, holding space, awareness of power dynamics, familiarity with non-ordinary states of consciousness, and offering a counterbalance to biomedical perspectives.^26^ For these reasons, we embedded SHCs as core members of our trial model of care.

### IV. Spiritually-Informed Triad Model

The overall protocol was modified to include a mental health clinician and spiritual health clinician dyad (as opposed to standard models, which typically include two mental health clinicians). Spiritual experiences are commonly reported in PAT and associated with improved treatment outcomes,^50^ and therapies incorporating spirituality in non-psychedelic formats have a strong empirical basis.^51^ As such, we modified standard supportive, non-directive psychedelic therapeutic model to include a “triad model” of care, emphasizing the patient, mental health clinician, and spiritual health clinician as vital members of the care team and therapeutic process, with the two clinicians using a collaborative approach to model trust, connection, and openness to the participant, and using a pluralistic, non-imposing, respectful approach that prioritized patient autonomy and empowerment that explored patient beliefs and worldviews (as it is important to note that all individuals have beliefs and worldviews, whether they include religion and spirituality or not). Both clinicians were present for all preparation sessions, the entire dosing day, and all integration sessions. Spiritual health clinicians are trained to support patients of any or no religious affiliation, and in many respects have more extensive training on spiritually competent care and non-imposition than those with solely mental health training, have been demonstrated to be effective at providing care in secular and pluralistic contexts, and have been recommended as clinicians for psychedelic therapies as well for this reason. Thus, spiritual health clinicians were part of the care team for all patients, regardless of religious affiliation.

Main goals of preparation sessions included facilitating therapeutic alliance and trust, practicing engaging in a non-judgmental attitude towards their internal experience, reviewing patient history and symptoms (including medical and mental health history as well as any SERT relevant, cultural factors, or belief systems), providing psychoeducation on the psilocybin session, exploring intentions, and practicing support strategies. During the psilocybin session, both clinicians provided a supportive presence and encouraged participant to engage with their internal experience in a nonjudgmental manner and provided specific support strategies when requested. During the integration sessions, both clinicians helped patients explore their psilocybin experience and engage in meaning making in a way that allowed the participant to interpret and understand their experience, with support from both clinicians, as well as discussing integration practices, or how to apply their insights or meaning to their daily life.

### V. Dosing Day Procedures

#### Description of Physical Environment

Dosing occurred in living room-like treatment room with a bed for the participant and two chairs for the facilitator dyad. The room was in a healthcare office building in a suburban environment near the Emory University main campus. The room had no windows. A book of landscapes was near the participant’s bed and images of nature were on the walls. The room had several synthetic plants, dim warm lighting, a fan, sound system, snacks, and other than the art on the walls, minimal additional decoration, though participants were welcomed to bring personal objects of significance to the room. The bed was at the far center of the room with the two chairs for the facilitators at either side of the foot of the bed. There was an additional ottoman flanking the bed for facilitators if they needed to approach the participant more closely. The bathroom also used dim lighting, and the bathroom mirror was covered with a print of a mountain. The private bathroom was ∼10 feet from the dosing room. Two doors down from the dosing room, a live feed of the dosing was reviewed by study coordinators. The study physician was also on site, also two doors down from the dosing room.

On the morning of the psilocybin treatment (Day 14), participants presented to the treatment area in the morning after eating a light, low-fat breakfast. They were expected to refrain from cannabinoid and as-needed opioid use the morning of dosing day. They were also asked to continue their basal opioid and/or long-acting benzodiazepine (if applicable) to avoid withdrawal. The day prior, patients received a urine drug screen, urine pregnancy test (if a woman of childbearing potential), and rapid testing for SARS-CoV-2. The day of dosing, participants again received a urine pregnancy test (if applicable). Participants were allowed to listen to their own music pre-dosing, and the study team prepared personalized playlists for participants if requested.

Participants also worked with their facilitators to curate a ritual of their choosing, if so desired (e.g., use of music, art, prayer). After participants were dosed with 25mg oral psilocybin, the standardized playlist was started (participants could not listen to their own music), and they were invited to review their intention for the treatment session, revisit grounding techniques addressed in preparatory psychoeducation, and engage in conversation with facilitators for up to 30 minutes. At the 30-minute mark, when the psychoactive effects of the psilocybin are expected to begin, participants were asked to lie down on a couch, wear eyeshades, and listen to preselected music during the session with headphones that was also played on a speaker system in the treatment room. Facilitators provided verbal reassurance for any psychologically challenging experiences and provided light, distal touch as consented by the patient prior to dosing. At around the six-hour mark, as the acute effects of the psilocybin were expected to wane, participants were invited to eat a light meal that was requested in advance by the participant. They were invited to write down their experience (or use other means of conveying their experience such as drawing) and to not discuss their experience in any detail with friends or family until after integration started the next day. At around the seven hour mark, the study physician evaluated the participant, and assessments pertaining to the acute psilocybin experience were administered around seven hours after dosing. Participants were released after the 8-hour mark after dosing to a trusted person whom the study team had met prior to dosing. Rescue medications (PO lorazepam and olanzapine) were available to treat any anxiety that could not be mitigated by facilitator support or to treat any psychiatric emergency. Dosing day occurred in a building with rapid response support to address any potential medical emergencies.

### VI. Heart Rate and Blood Pressure Figures

**Figure 1.**
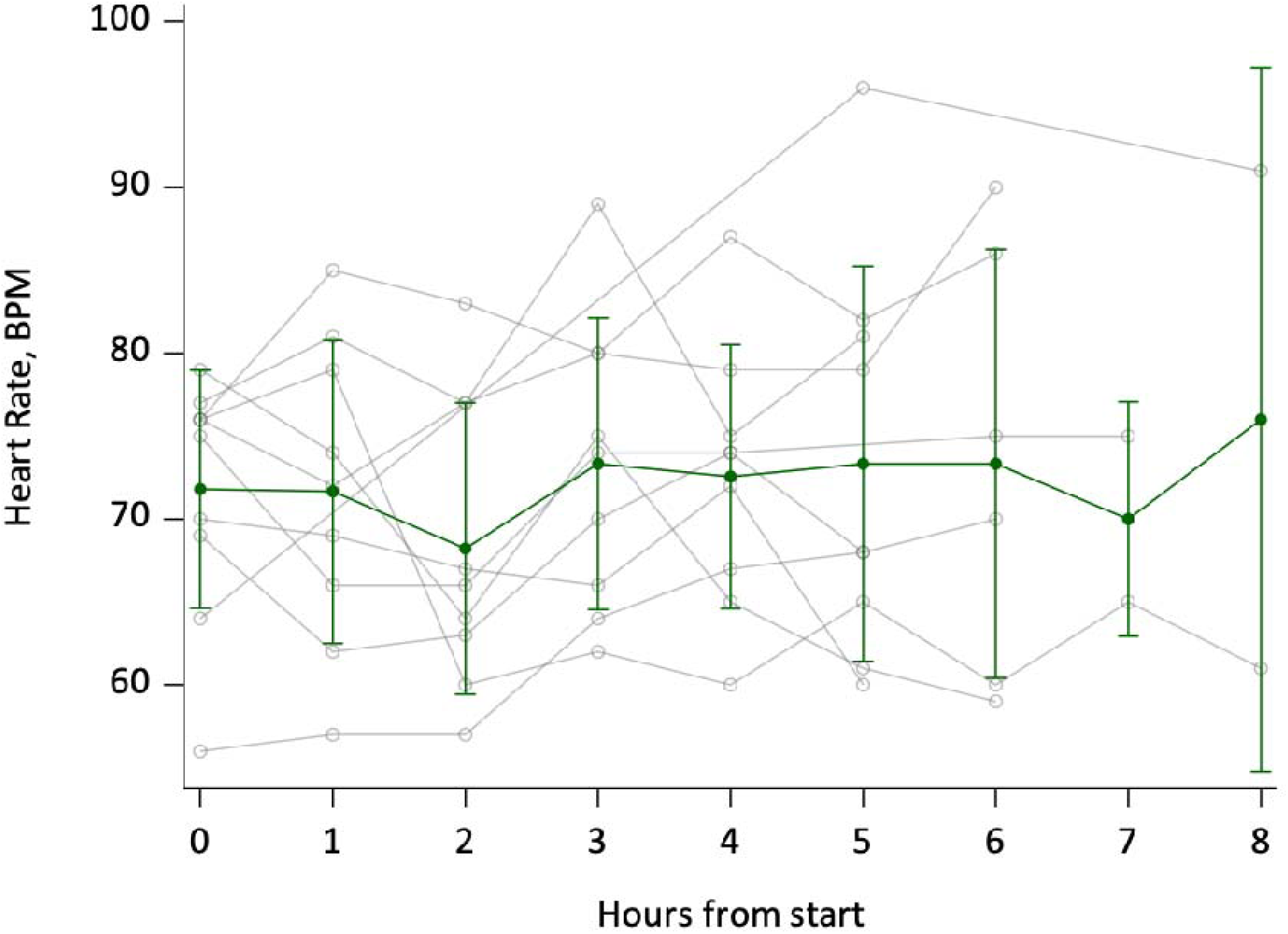
Hourly heart rate (BPM). Error bars are 1 SD.

**Figure 2.**
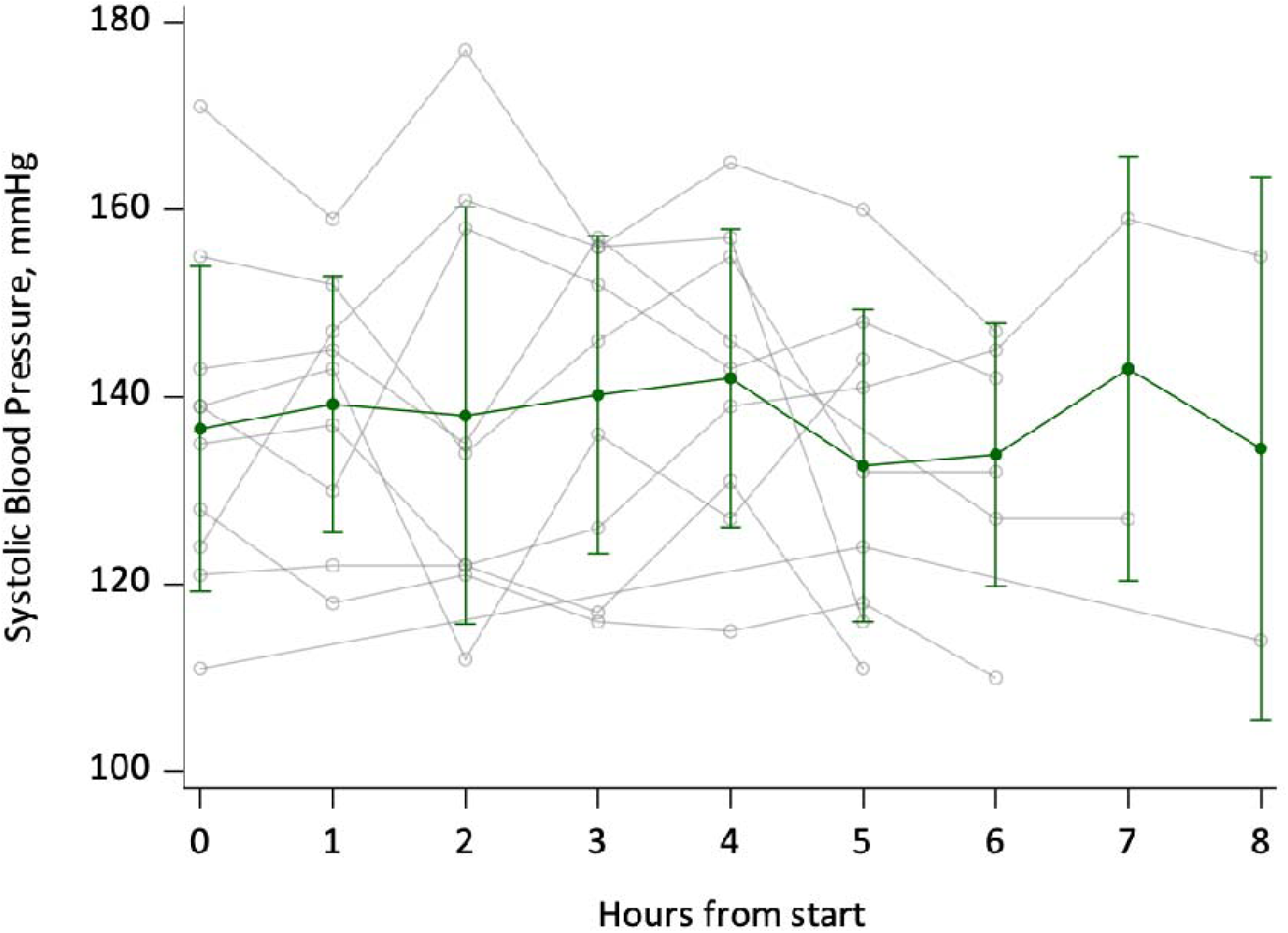
Systolic blood pressure, mmHg. Error bars are 1 SD.

**Figure 3.**
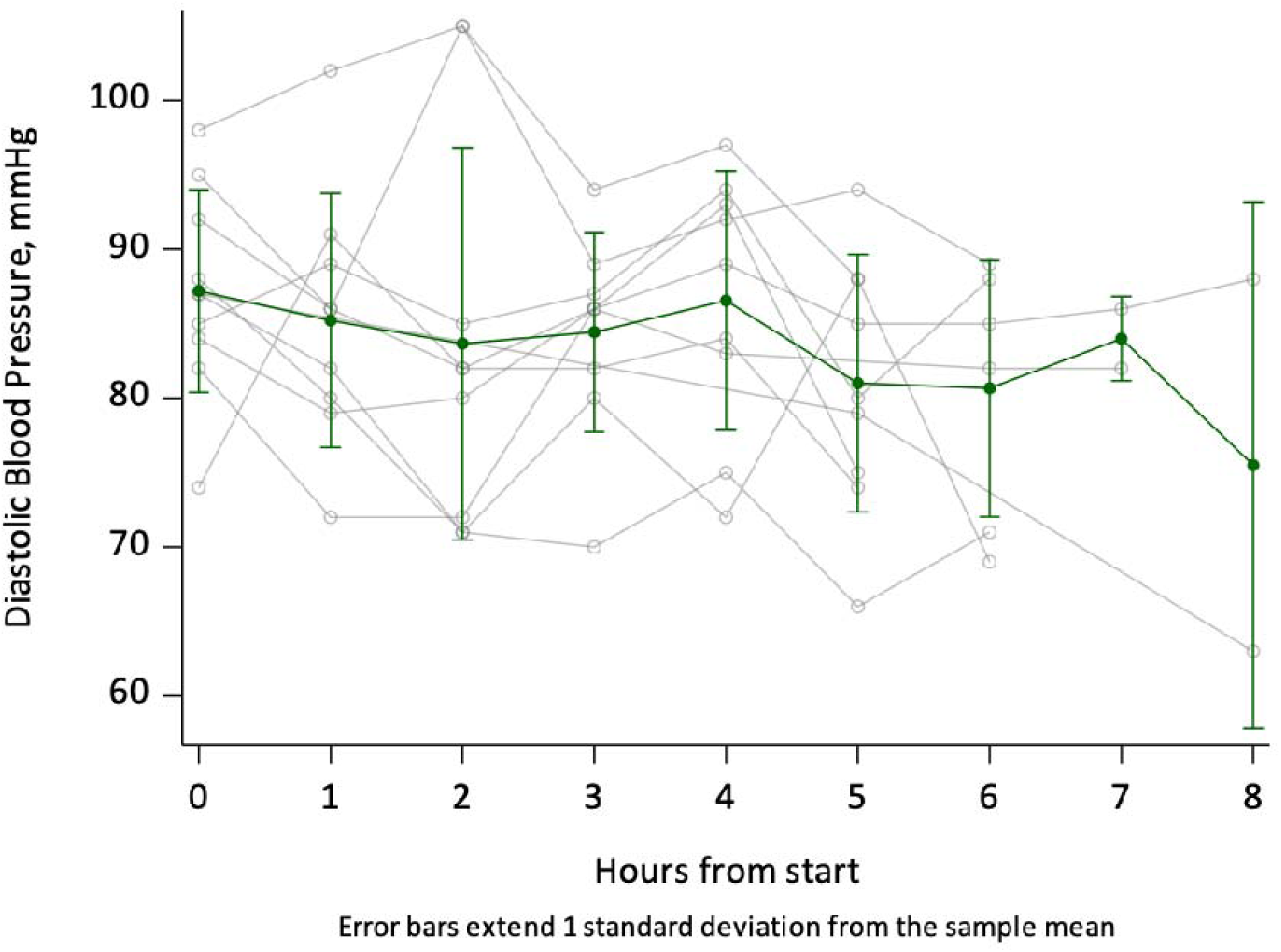
Diastolic blood pressure, mmHg. Error bars are 1 SD.

### VII. Complete Table of Exploratory Outcomes and Assessments

| Exploratory Outcomes and Assessments |  |
| --- | --- |
| *Participant outcomes and assessments* |  |
| Outcomes | Assessments |
| Demoralization | Demoralization Scale (DSII) |
| Pain intensity and interference | Brief Pain Inventory (BPI) |
| Pain Catastrophizing | Pain Catastrophizing Scale (PCS) |
| Quality-of-Life | Harvard Flourishing Measure (HFM) |
| Anxiety and Depression | Hospital Anxiety and Depression Scale (HADS) |
| Global symptom burden | Edmonton Symptom Assessment Scale (ESAS) |
| Global psychosocial functioning | Work and Social Adjustment Scale (WSAS) |
| Mystical-Type Experience | Mystical Experience Questionnaire (MEQ) |
| Challenging experiences related to psychedelics | Challenging Experience Questionnaire (CEQ) |
| Strength of religious faith and engagement | Santa Clara Strength of Religious Faith Questionnaire (SCSORF) |
| Measure of awe emotion | Awe Experience Scale (AWE-S) |
| Emotional breakthroughs related to psychedelic experience | Emotional Breakthrough Inventory (EBI) |
| Post-traumatic Growth | Post-Traumatic Growth Inventory-Short Form (PTGI-SF) |
| Social connectedness | Social Connectedness Scale-Revised (SCS-R) |
| Opioid use | Oral morphine equivalents (OME) |
| Overall experience of life stressors | National Institutes of Health – Healing Experience of All Life Stressors (NIH-HEALS) |
| Fear of cancer recurrence* | Fear of Cancer Recurrence Inventory-Short Form (FCRI-SF) |
| *Caregiver outcomes and assessments* |  |
| Global psychosocial functioning | Work and Social Adjustment Scale (WSAS) |
| Caregiver strain | Modified Caregiver Strain Index (MCSI) |

\*Only administered to participants in remission or no active disease

### VIII. Complete Table of Adverse Events

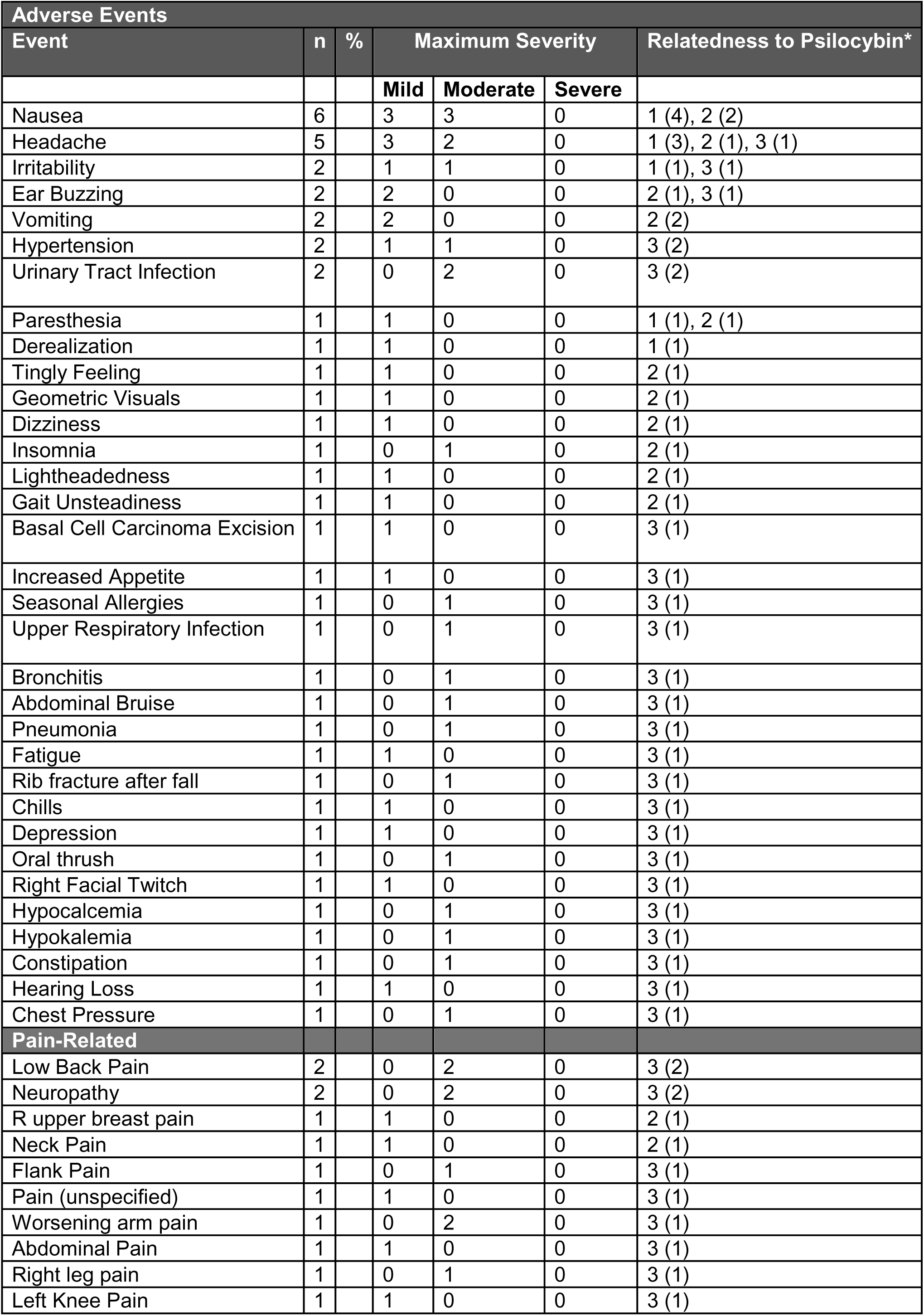

| Adverse Events |  |  |  |  |  |  |
| --- | --- | --- | --- | --- | --- | --- |
| Event | n | % | Maximum Severity |  |  | Relatedness to Psilocybin* |
|  |  |  | Mild | Moderate | Severe |  |
| Nausea | 6 |  | 3 | 3 | 0 | 1 (4), 2 (2) |
| Headache | 5 |  | 3 | 2 | 0 | 1 (3), 2 (1), 3 (1) |
| Irritability | 2 |  | 1 | 1 | 0 | 1 (1), 3 (1) |
| Ear Buzzing | 2 |  | 2 | 0 | 0 | 2 (1), 3 (1) |
| Vomiting | 2 |  | 2 | 0 | 0 | 2 (2) |
| Hypertension | 2 |  | 1 | 1 | 0 | 3 (2) |
| Urinary Tract Infection | 2 |  | 0 | 2 | 0 | 3 (2) |
| Paresthesia | 1 |  | 1 | 0 | 0 | 1 (1), 2 (1) |
| Derealization | 1 |  | 1 | 0 | 0 | 1 (1) |
| Tingly Feeling | 1 |  | 1 | 0 | 0 | 2 (1) |
| Geometric Visuals | 1 |  | 1 | 0 | 0 | 2 (1) |
| Dizziness | 1 |  | 1 | 0 | 0 | 2 (1) |
| Insomnia | 1 |  | 0 | 1 | 0 | 2 (1) |
| Lightheadedness | 1 |  | 1 | 0 | 0 | 2 (1) |
| Gait Unsteadiness | 1 |  | 1 | 0 | 0 | 2 (1) |
| Basal Cell Carcinoma Excision | 1 |  | 1 | 0 | 0 | 3 (1) |
| Increased Appetite | 1 |  | 1 | 0 | 0 | 3 (1) |
| Seasonal Allergies | 1 |  | 0 | 1 | 0 | 3 (1) |
| Upper Respiratory Infection | 1 |  | 0 | 1 | 0 | 3 (1) |
| Bronchitis | 1 |  | 0 | 1 | 0 | 3 (1) |
| Abdominal Bruise | 1 |  | 0 | 1 | 0 | 3 (1) |
| Pneumonia | 1 |  | 0 | 1 | 0 | 3 (1) |
| Fatigue | 1 |  | 1 | 0 | 0 | 3 (1) |
| Rib fracture after fall | 1 |  | 0 | 1 | 0 | 3 (1) |
| Chills | 1 |  | 1 | 0 | 0 | 3 (1) |
| Depression | 1 |  | 1 | 0 | 0 | 3 (1) |
| Oral thrush | 1 |  | 0 | 1 | 0 | 3 (1) |
| Right Facial Twitch | 1 |  | 1 | 0 | 0 | 3 (1) |
| Hypocalcemia | 1 |  | 0 | 1 | 0 | 3 (1) |
| Hypokalemia | 1 |  | 0 | 1 | 0 | 3 (1) |
| Constipation | 1 |  | 0 | 1 | 0 | 3 (1) |
| Hearing Loss | 1 |  | 1 | 0 | 0 | 3 (1) |
| Chest Pressure | 1 |  | 0 | 1 | 0 | 3 (1) |
| Pain-Related |  |  |  |  |  |  |
| Low Back Pain | 2 |  | 0 | 2 | 0 | 3 (2) |
| Neuropathy | 2 |  | 0 | 2 | 0 | 3 (2) |
| R upper breast pain | 1 |  | 1 | 0 | 0 | 2 (1) |
| Neck Pain | 1 |  | 1 | 0 | 0 | 2 (1) |
| Flank Pain | 1 |  | 0 | 1 | 0 | 3 (1) |
| Pain (unspecified) | 1 |  | 1 | 0 | 0 | 3 (1) |
| Worsening arm pain | 1 |  | 0 | 2 | 0 | 3 (1) |
| Abdominal Pain | 1 |  | 1 | 0 | 0 | 3 (1) |
| Right leg pain | 1 |  | 0 | 1 | 0 | 3 (1) |
| Left Knee Pain | 1 |  | 1 | 0 | 0 | 3 (1) |

### IX. Retrospective Questionnaire Assessing meaning, educational, and spiritual impacts of the intervention

Responses to the following prompts:

*Meaning: How personally meaningful were the experiences in your medication session? Educational: How educational were the experiences in your medication session?*

*Spiritual: Indicate the degree to which the experiences in your medication session were spiritually significant to you*.

Likert scale responses: not at all, slightly, moderately, very much, among the top 5 most spiritually significant experiences of your life, the single most spiritually significant experience of my life.

| Participant Number | Meaning | Educational | Spiritual |
| --- | --- | --- | --- |
| 01 | Among the 5 most meaningful experiences of my life | Among the 5 most meaningful experiences of my life | Among the 5 most spiritual experiences of my life |
| 02 | The single most meaningful experience of my life | The single most educational experience of my life | Among the 5 most spiritual experiences of my life |
| 03 | Among the 5 most meaningful experiences of my life | The single most educational experience of my life | The single most spiritual experience of my life |
| 04 | Similar to meaningful experiences that occur on average once every 5 years | Similar to meaningful experiences that occur on average once every 5 years | Slightly |
| 05 | Similar to meaningful experiences that occur on average once a month | Among top 10 meaningful experiences of my life | Slightly |
| 06 | Among the 5 most meaningful experiences of my life | Among the 5 most meaningful experiences of my life | Among the 5 most meaningful experiences of my life |
| 07 | Among the 5 most meaningful experiences of my life | Among the 5 most meaningful experiences of my life | Among the 5 most meaningful experiences of my life |
| 08 | Among the 5 most meaningful experiences of my life | Among the 5 most meaningful experiences of my life | Among the 5 most meaningful experiences of my life |
| 09 | Among the 5 most meaningful experiences of my life | Among the 5 most meaningful experiences of my life | Very much |
| 10 | Similar to meaningful experiences that occur on average once a year | Similar to meaningful experiences that occur on average once a year | Slightly |

### X. Pain Catastrophizing Subscale Domains – Table and Figures

| Pain Catastrophizing Subscales |  |  |  |  |  |
| --- | --- | --- | --- | --- | --- |
|  |  | Raw Score |  | Change from Baseline |  |
| Outcome | Time | N | Median (IQR) | N | Median (IQR) |
| **Pain Catastrophizing – Magnification** |  |  |  |  |  |
|  | D0 | 10 | 6 (5, 8) | 10 | 0 (0, 0) |
|  | D13 | 10 | 5 (4, 8) | 10 | -0.5 (-2, 2) |
|  | D15 | 10 | 2 (1, 3) | 10 | -3.5 (-6, -2) |
|  | D42 | 10 | 0 (0, 2) | 10 | -4.5 (-7, -2) |
|  | D98 | 10 | 1.5 (0, 3) | 10 | -4.0 (-6, -2) |
| **Pain Catastrophizing – Rumination** |  |  |  |  |  |
|  | D0 | 10 | 8.5 (5, 12) | 10 | 0 (0, 0) |
|  | D13 | 10 | 11.5 (8, 12) | 10 | 1 (-1, 2) |
|  | D15 | 10 | 6 (0, 9) | 10 | -3.5 (-10, 4) |
|  | D42 | 10 | 2.5 (0, 5) | 10 | -4.5 (-11, 0) |
|  | D98 | 10 | 2.5 (0, 12) | 10 | -4.5 (-11, 3) |
| **Pain Catastrophizing - Helplessness** |  |  |  |  |  |
|  | D0 | 10 | 12 (5, 15) | 9 | 0 (0, 0) |
|  | D13 | 10 | 12 (7, 15) | 9 | -0.5 (-3, 2) |
|  | D15 | 10 | 4 (0, 7) | 9 | -5.5 (-15, -1) |
|  | D42 | 10 | 1.5 (0, 5) | 9 | -9.5 (-13, -1) |
|  | D98 | 10 | 3.5 (0, 7) | 9 | -7.0 (-14, -1) |

**Figure 1.**
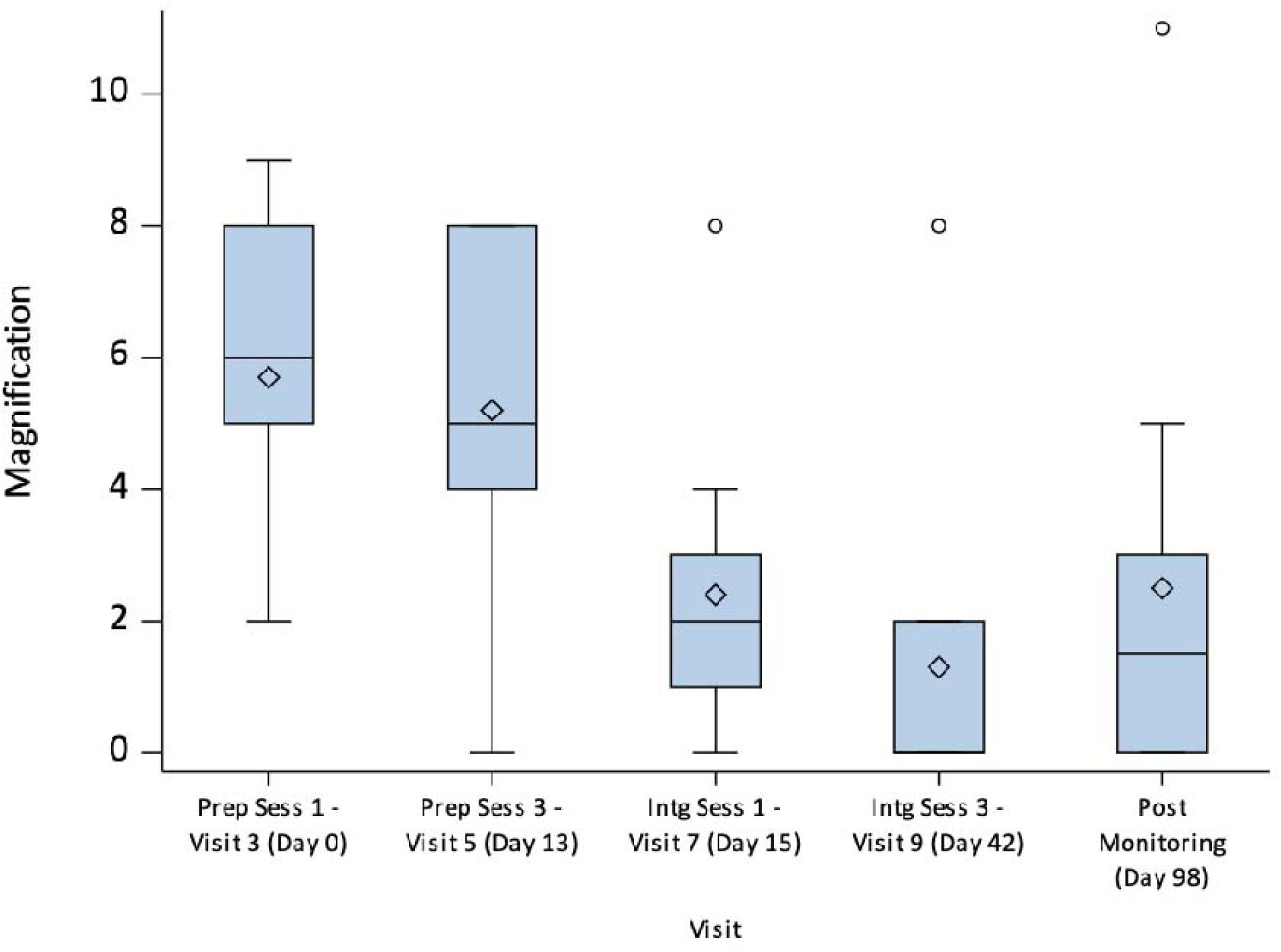
Pain Catastrophizing – Magnification scores from baseline to day 98 with box plots representing IQR.

**Figure 2.**
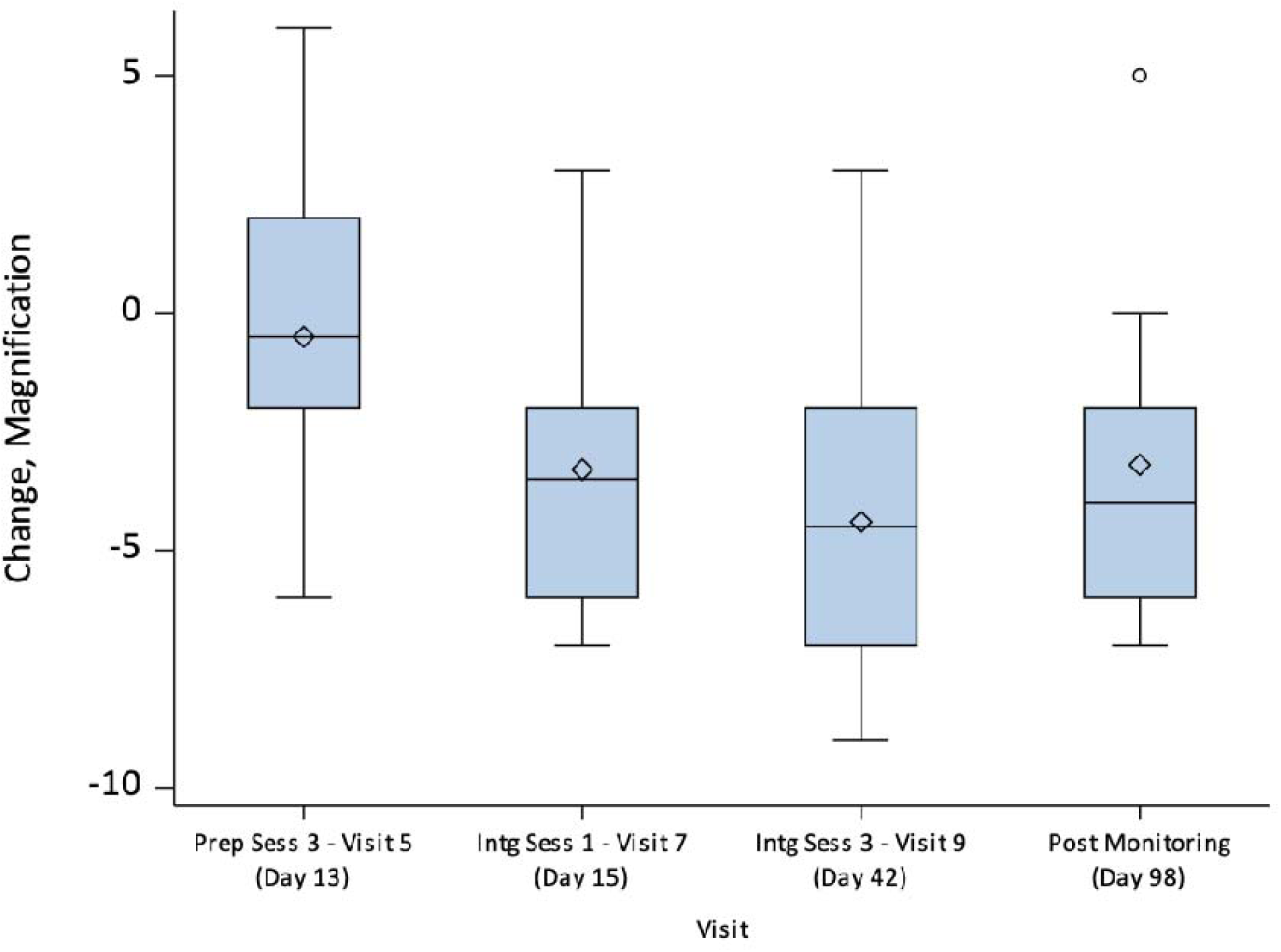
Change from baseline in Pain Catastrophizing – Magnification scores from baseline to day 98 with box plots representing IQR.

**Figure 3.**
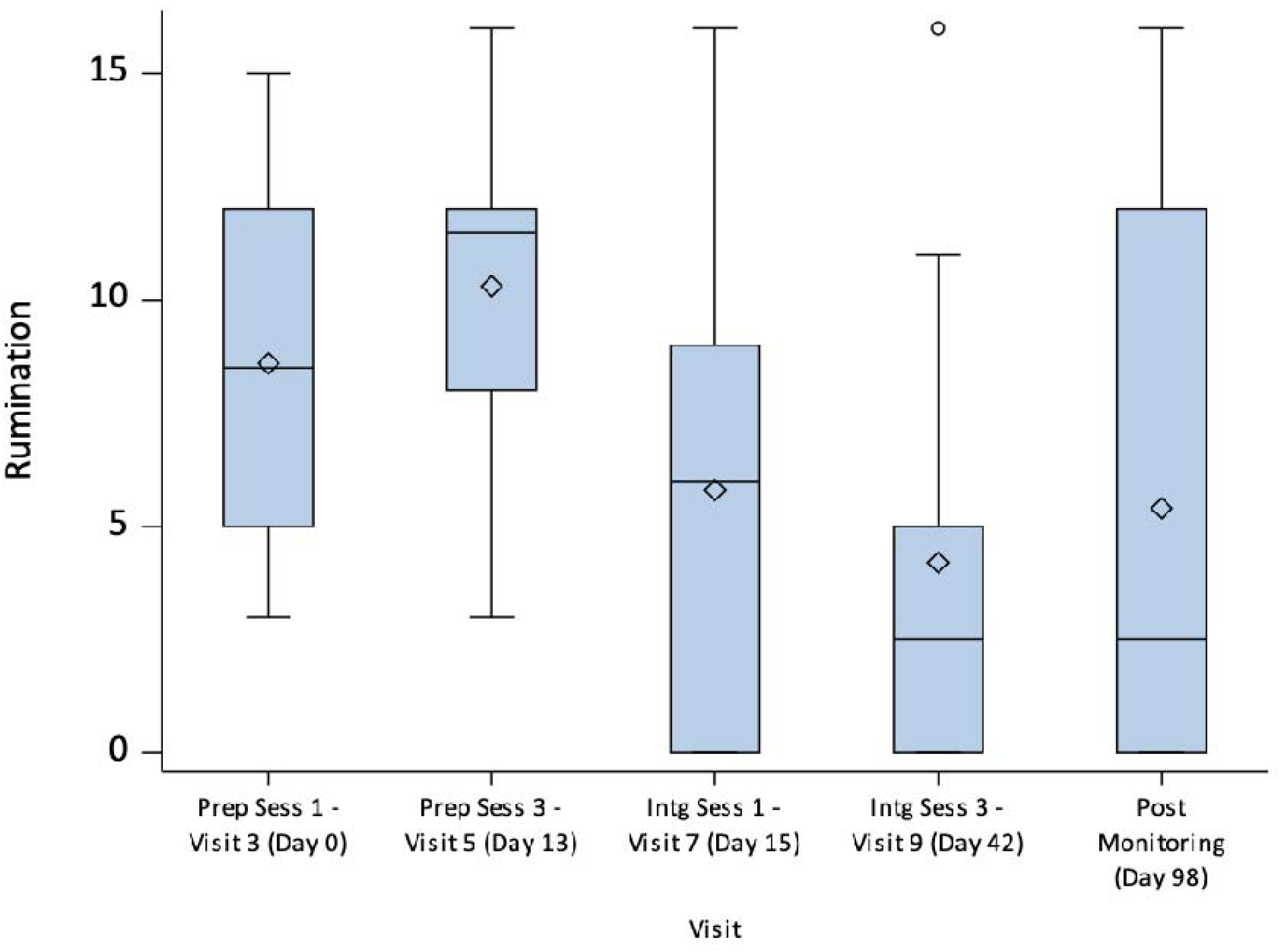
Pain Catastrophizing –Rumination scores from baseline to day 98 with box plots representing IQR.

**Figure 4.**
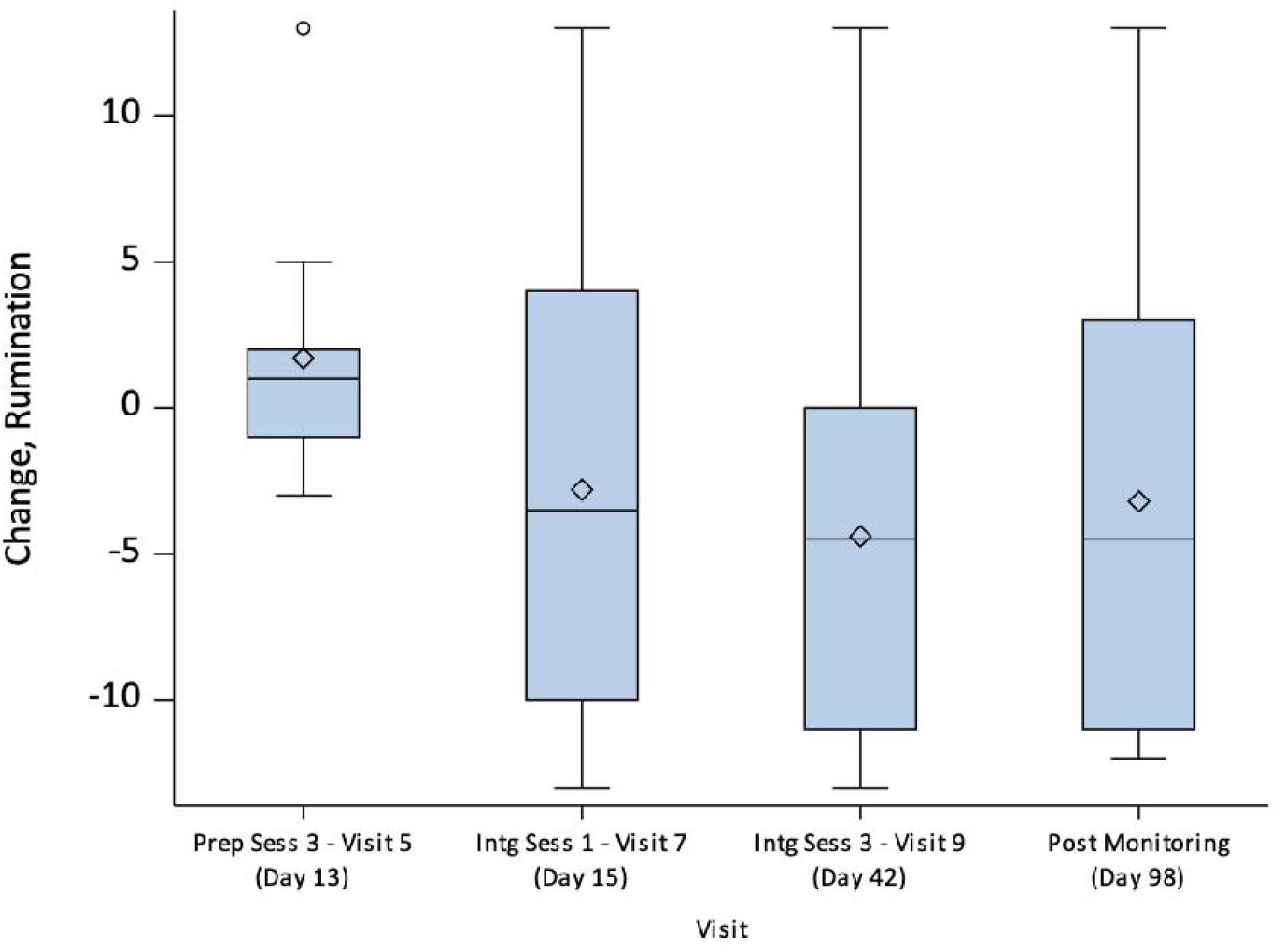
Change from baseline in Pain Catastrophizing – Rumination scores from baseline to day 98 with box plots representing IQR.

**Figure 5.**
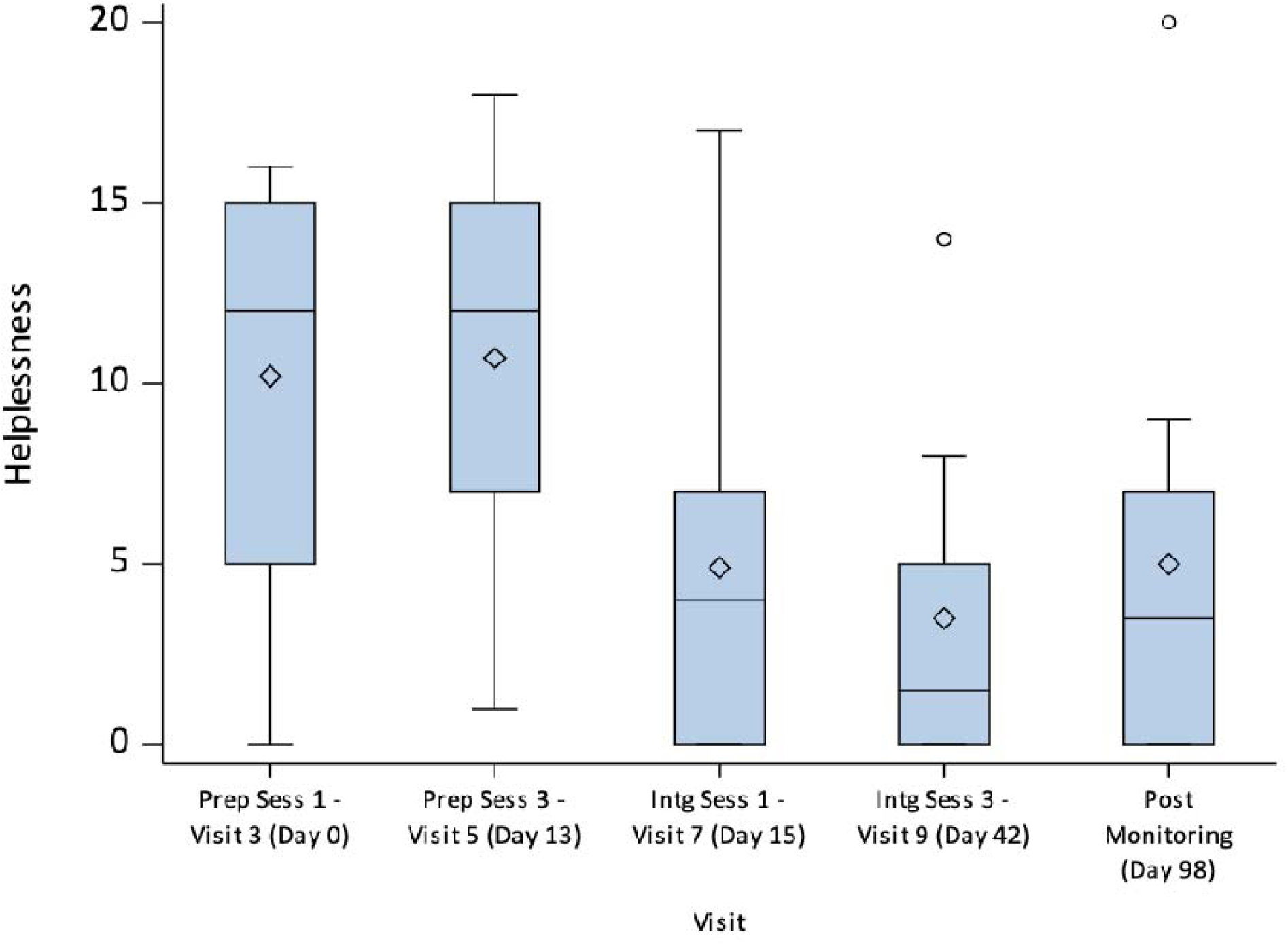
Change in Pain Catastrophizing – Helplessness scores from baseline to day 98 with box plots representing IQR.

**Figure 6.**
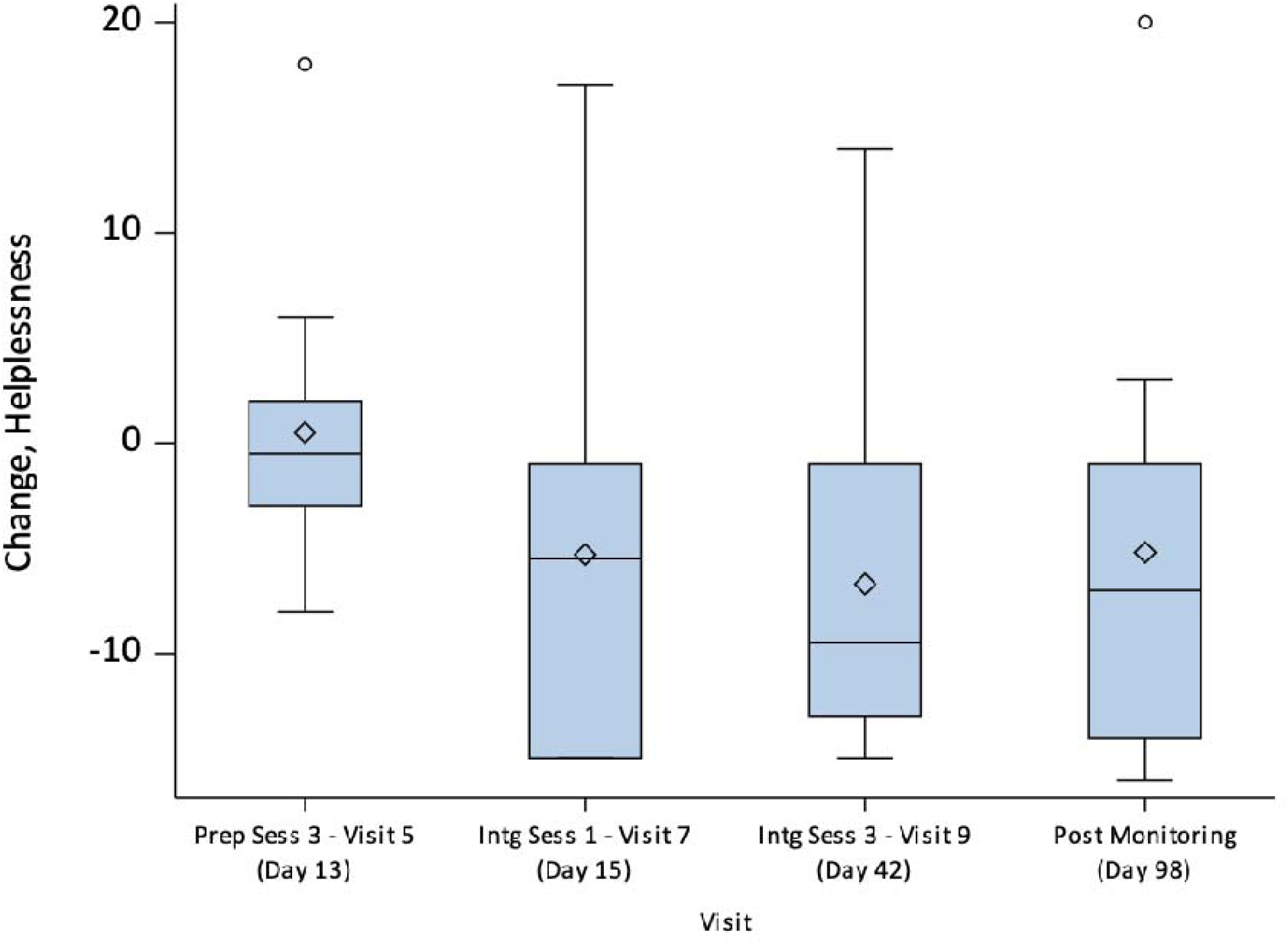
Change from baseline in Pain Catastrophizing – Helplessness scores from baseline to day 98 with box plots representing IQR.

### XI. Life Satisfaction Figure

*Life satisfaction score at baseline, Day 42 (study endpoint), and 98 presented as box plots with interquartile range (IQR). Dashed line represents the median and diamond represents the mean*.

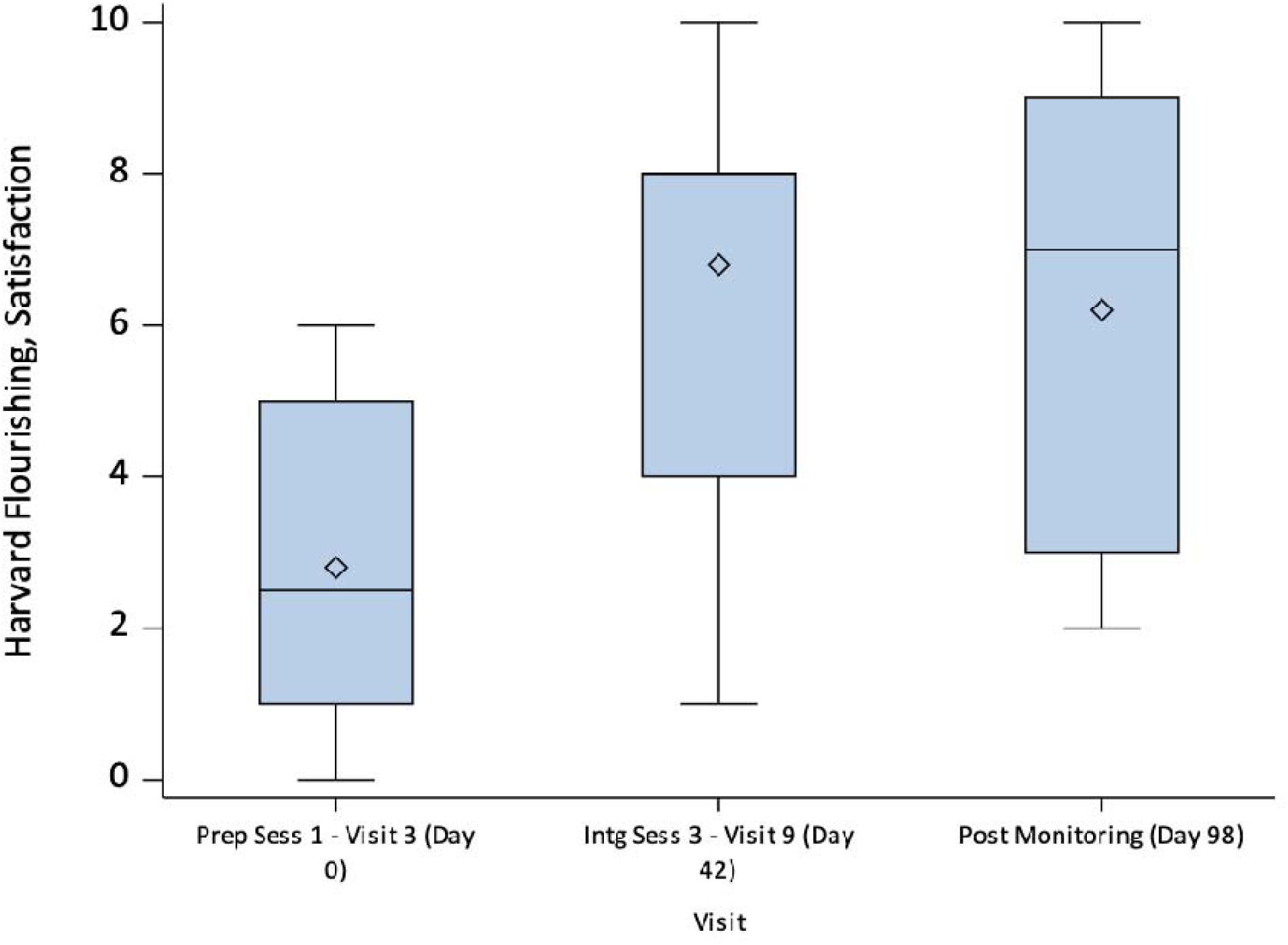

### XII. Narratives of the Dosing Experience from the Retrospective Questionnaire

Representative narratives in response to the prompt: *please briefly describe what was most memorable, meaningful, educational, and/or spiritually significant about the experiences in your medication session*.

#### Sense of Connectedness

“The sense of connectedness I felt during the psilocybin experience was unlike anything I’d felt before. I also had a tremendous sense of wellbeing and wholeness of my body that I haven’t experienced before. [My facilitators] made the experience safe and comfortable, and truly helped make the experience transformative for me”

“I have within me what I need to face and accept fear, pain, cancer, etc. It’s been there the whole time, I just lost sight of it. The connectedness I found showed me the foundation of the life I’ve built and created for my family and loved ones.”

“Increased appreciation for how connected we all are.”

“[I feel] more fragmented but more cohesive and connected.”

#### Sense of the Sacred

“I felt the presence of God – He held His hand out to me. But I had the sense that it wasn’t time for me to take it yet. But He’ll be there when it is.”

“The medicine and God worked together to show me the purpose of my life. I learned real forgiveness regarding my cancer. I know I’m not alone and that my life has had a purpose. All of my experiences felt integrated and purposeful.

“In session I received confirmation of spiritual awakening. I have been on a spiritual journey with lots of questions and doubts. This session confirmed to me I was on the right path. Keep going.”

#### Sense of Gratitude

“I felt that I am a new person entirely, but realize it is a work in progress. I am extremely grateful for this opportunity. I truly believe it has been life changing and will extend my life.”

“I felt gratitude from G-d to fulfilling my purpose in life.”

#### Transcendence from Pain

“I was able to see past the hurt and pain inside and was met with love. Unconditional love. Love for self and others…I feared the cancer returning, and the thought of my pain never going away. I now feel empowered.”

“[I feel] a stronger desire to fight pain or fatigue to be a benefit to those I love.”

“I’m much less irritable. Pain is significantly reduced. Considerably more accepting of myself and the trauma I’ve experienced.”

“My brain is rewired. It changed my life. I have more wellbeing, peace, joy. Significantly less pain.”

“I have never had any experience similar to this. Only once, when a very kind person offered to pray with me during a difficult time did I feel something somewhat of this nature. Then I felt the pain lift and a golden light flood me. I was completely surprised.”

### XIII. Consent Procedures

Participants were made aware of common adverse events related to psilocybin, including the following:

#### Common (over 50%)

1. Visual and other sensory distortions, feeling of unreality & changed sense of time
2. Anxiety at the onset of the drug effects
3. Increased heart rate and blood pressure (We will check your heart function at screening and patients with heart abnormalities will be excluded from the study)

#### Less common (about 10-40%)

1. Nausea
2. Dizziness, blurred vision, drowsiness & sleepiness
3. Headache
4. Temporary suspiciousness

#### Rare (< 10%)

1. “Flashbacks”, or hallucinogen-induced persistent perceptual disorder (HPPD). This adverse effect is seen rarely following recreational use and has not been reported in scientific studies done under supportive clinical conditions
2. Worsening of mental state after the drug experience (very rare and was not seen in similar studies).

*The full informed consent form can be provided upon request*.

### XIV. ReSPCT Guidelines 2025 – Reporting Table

Pronovost-Morgan C, Greenway KT, Roseman L, The ReSPCT Experts. An international Delphi consensus for reporting of setting in psychedelic clinical trials. *Nat Med.* 2025.

https://doi.org/10.1038/s41591-025-03685-9

| Item | Item Number | Item details | Reported on page number: |
| --- | --- | --- | --- |
| **Physical environment** |  |  |  |
| Location | 1 | Location of the trial, and whether indoors/outdoors and urban/rural/suburban. | 24 |
| Ambiance | 2 | Room ambiance curated by the study team. | 24 |
| Access to nature | 3 | Sources of nature or natural elements that are physically or visually accessible to participants. | 24 |
| Objects and decorations | 4 | Objects and decorations in the room. | 24 |
| Lighting | 5 | Room lighting and adjustability. | 24 |
| Sensory reduction | 6 | Sensory reduction used, such as headphones and eyeshades. | 24 |
| Bathroom facilities | 7 | Level of bathroom accessibility and privacy. | 24 |
| **Dosing session procedure** |  |  |  |
| Number and roles of people present | 8 | Number and roles of people present, including participants, study staff and informal support. | 24 |
| Positioning | 9 | Relative position of people in the room, and what participants were positioned on (e.g., bed, mat, couch). | 24 |
| Focus and main activities | 10 | Focus (internal or external) and main activities of the dosing session. | 24 |
| Music or soundscapes | 11 | Music or soundscapes that accompanied the dosing experience. | 24 |
| Interpersonal interventions | 12 | Verbal or physical interpersonal interventions used throughout the session, and how consent was obtained. | 24 |
| Participant autonomy, control, and agency | 13 | Level of participant control and agency over activities and environment of the dosing session. | 4, 23 |
| Dosing regimen | 14 | Dosing regimen, including drug dose(s), frequency, route of administration and length of the. | 3-4 |
| Medical and experimental procedures or assessments | 15 | Medical and experimental procedures or assessments performed during the dosing session. | 40 |
| Pre- and post-dosing protocol | 16 | Activities that took place immediately prior to- or post-dosing, including participant arrival and release conditions. | 24 |
| Potential disturbances or interruptions | 17 | Disturbances or interruptions that may have impacted the quality of the dosing session. | Qualitative Paper (citation 48) |
| **Therapeutic framework and protocol** |  |  |  |
| Therapeutic or guiding approach | 18 | Therapeutic/guiding approach used throughout the study, if any, with the accompanying manual or protocol. | 26-27 |
| Narrative framing | 19 | Framing of the trial intervention by the study team, including the short- and long-term drug effects. | 24 |
| Number of sessions | 20 | Number and length of preparation, dosing, and integration sessions. | 3-4 |
| Preparation protocol | 21 | Activities performed during the preparation sessions. | 3-4 |
| Integration protocol | 22 | Activities performed during the integration sessions. | 3-4 |
| Additional support/follow-up | 23 | Formal or informal support or follow-up offered to participants after the end of the trial. | 3 |
| Study personnel qualifications | 24 | The credentials, training, and expertise of personnel providing the study intervention or care. | 24 |
| Cultural competence and safety | 25 | Study team's level of cultural competence and efforts towards cultural safety. | 24 |
| **Subjective experiences** |  |  |  |
| Therapeutic alliance | 26 | Therapeutic alliance between participants and facilitators throughout the intervention. | Qualitative Paper (citation 48) |
| Trust | 27 | Participant's level of trust throughout the intervention. | Qualitative Paper (citation 48) |
| Physical comfort | 28 | Participant's level of physical comfort during the dosing session. | Qualitative Paper (citation 48) |
| Physical safety | 29 | Participant's sense of physical safety during the dosing session. | Qualitative Paper (citation 48) |
| Psychological and cultural safety | 30 | Participant's sense of interpersonal safety with the people present during the session. | Qualitative Paper (citation 48) |

### XV. Dosing Room Image

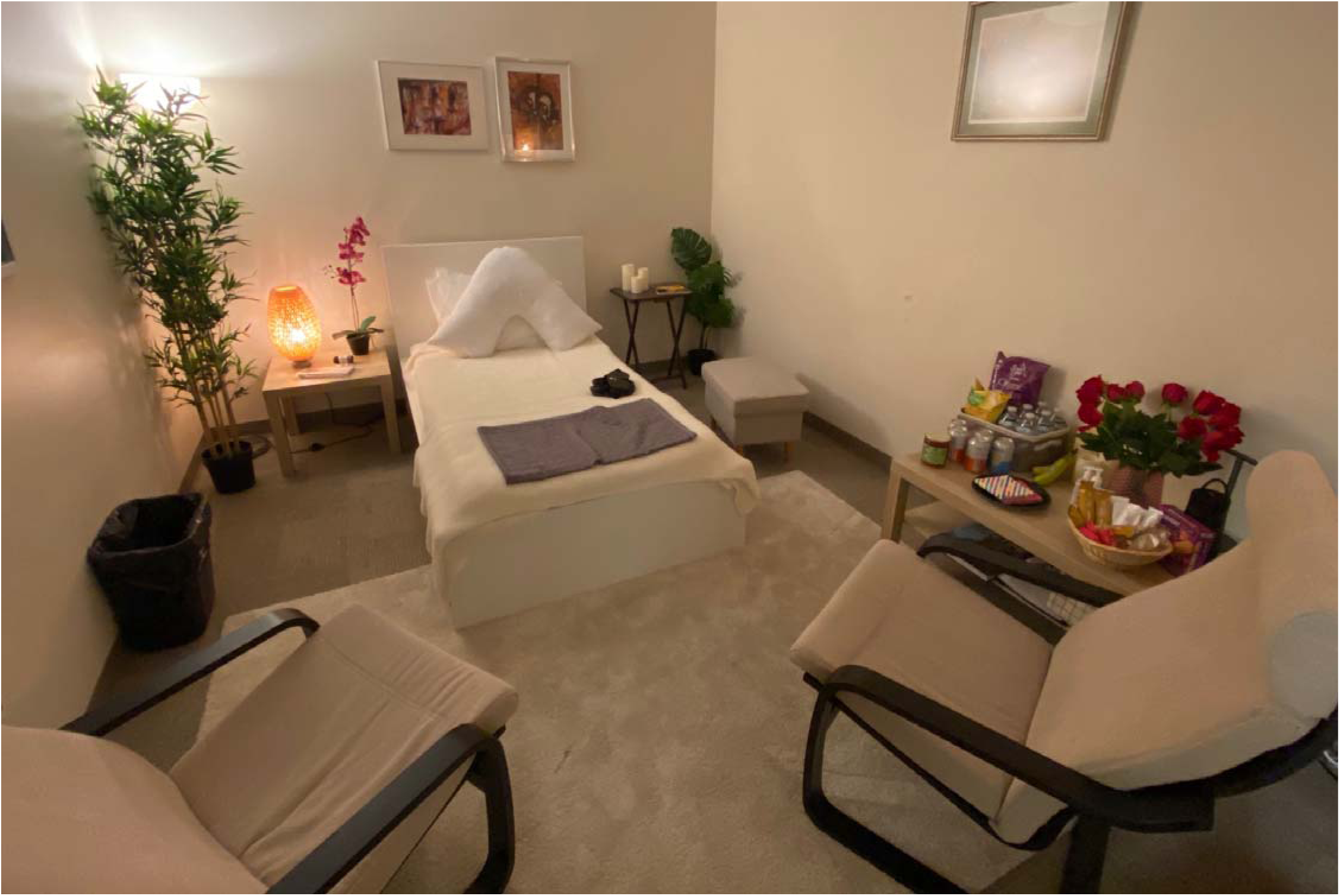

## Notes

### Competing Interest Statement

Ali J. Zarrabi
Disclosures: Dr. Zarrabi has received research support from Compass Pathways, Reunion Neuroscience, and the Usona Institute.
Boadie W. Dunlop
Disclosures: Dr. Dunlop has received research support from Beckley PsyTech, Boehringer Ingelheim, Compass Pathways, LivaNova, NIMH, Reunion Neuroscience, Usona Institute and served as a consultant to Myriad Neuroscience and Otsuka.
Barbara O. Rothbaum
Disclosures: Dr. Rothbaum has been paid as a consultant by Otsuka, Psychwire, Senseye, MAPS, Lykos, Jazz Pharmaceuticals, GoodCap, Transcend, Penumbra.
Roman Palitsky
Disclosures: Dr. Palitsky has received research funding from the Tiny Blue Dot Foundation, the Riverstyx Foundation, Shefa, the Common Era Fund, and the Vail Health Foundation. He is a consultant for projects conducted at Harvard Divinity School.
Charles Raison:
Disclosures: Dr. Raison serves as a consultant for Usona Institute, Eli Lilly, Otsuka and AbbVie and receives research funding from the Tiny Blue Dot Foundation, Vail Health Foundation and ARPA-H.
Jessica Maples-Keller
Disclosures: Dr. Maples-Keller has received research funding from the Department of Defense, the state of Georgia, and the Wounded Warrior Project, and Multidisciplinary Association of Psychedelic studies, and has received consulting payments from Compass Pathways and Otsuka.

### Clinical Trial

NCT05506982

